# Nulliparity modifies the maternal genetic effects on gestational duration but not on birth weight

**DOI:** 10.1101/2025.06.17.25329777

**Authors:** Karin Ytterberg, Hedvig Sundelin, Xiaoping Wu, Julius Juodakis, Marc Vaudel, Elizabeth C. Corfield, Ole A. Andreassen, Per Magnus, Alexandra Havdahl, Staffan Nilsson, Pål R. Njølstad, Stefan Johansson, Bo Jacobsson, Pol Solé-Navais

## Abstract

Gestational duration and birth weight are linked to adverse health outcomes in offspring and mothers. Previous genome-wide association studies (GWAS) on these pregnancy outcomes did not consider parity, despite its known effects on mean values and variance. Here, we explored whether nulliparity (no previous births) modified the maternal or fetal genetic effects on gestational duration and birth weight, and resolved effect origin in up to 58,528 mothers and fetuses from the Norwegian Mother, Father, and Child Cohort Study. We identified shared and distinct loci for each outcome in parity-stratified GWAS. For gestational duration, we identified more maternal loci in nulliparous women, and a polygenic predictor had a three-fold higher prediction accuracy among nulliparous compared to multiparous women (p-value interaction = 5×10^−5^), with validation in an independent cohort. The results for birth weight showed discrepancies between the number of loci and the polygenic predictor. Our study reveals that nulliparity modifies the maternal genetic effects on gestational duration and highlights the relevance of considering nulliparity in genetic, and likely experimental, studies on pregnancy outcomes.

**Teaser:** Genome-wide study highlights that maternal genetics affect pregnancy length more strongly in first pregnancy.

## Introduction

The duration of gestation and offspring’s birth weight are linked to short- and long-term adverse health outcomes in both the mother and the neonate. For instance, preterm birth (before 37 weeks of gestation) is the leading cause of death in neonates and children under five years of age (1). Gestational duration and birth weight are complex traits with polygenic architectures; previous genome-wide association studies (GWASs) have identified multiple genetic loci associated with these birth outcomes in both the maternal and fetal genomes (n up to 321,223 across studies) (2–6). While the maternal genome has a larger role in shaping the duration of gestation compared to the fetal one, it’s the opposite for birth weight; the fetal genome contributes more to its genetic architecture than the maternal one. Overall, 30 and 250 loci have been identified to date for gestational duration and birth weight, respectively. However, these GWASs have overlooked information on the number of previous pregnancies of a mother.

Parity, the number of children a mother has previously delivered, is an important clinical factor. Today, we know that gestational duration is shortest and has higher variance in the first pregnancy leading to a birth (nulliparous women) (7), and that birth weight (8, 9) also tends to be lower in nulliparous compared to multiparous women (i.e., women who have given birth at least once). The immune system (10, 11) and uterine size (12) differ by parity and may potentially contribute to the differences observed in the mean and distributions of these pregnancy outcomes. These mechanisms could also play a role in modulating the genetic effects of the maternal and fetal genomes on the duration of gestation (7) and birth weight, but this has been largely ignored in GWASs performed to date.

The classical approach to testing gene × environment interaction effects is to investigate each genetic variant separately (13–15), which has a high multiple testing burden. To increase power, different polygenic approaches have been proposed, including investigating potential heritability differences or estimating genetic correlations between the different environmental groups (16–18). An alternative is to assess whether the environmental factor interacts with the cumulative effect of thousands of genetic variants across the genome, represented as a polygenic score (PGS) (18–21).

In this study we investigated whether nulliparity modifies the maternal or fetal genetic effects on gestational duration and birth weight in up to 64,000 mothers/ children. We explored parity-stratified GWASs in pregnancies of nulliparous and multiparous women, single nucleotide polymorphism (SNP) × nulliparity interactions, if the polygenic prediction of any outcome interacted with nulliparity, and lastly the genetic correlation between pregnancies of nulliparous and multiparous women. Our results highlight that nulliparity modifies the maternal genetic effect on gestational duration, but for birth weight modification of genetic effects were more inconsistent.

## Results

### Parity-stratified analyses indicate different lead variants by parity

First, we explored the differences in the number of lead loci identified in parity-stratified maternal and fetal GWAS of gestational duration and birth weight using the the Norwegian Mother, Father, and Child Cohort Study (MoBa) cohort (Table S1). We also assessed the effect origin (maternal or fetal) of lead SNPs by using haplotype-based analyses of the maternal transmitted and non-transmitted and paternal transmitted alleles. The distributions of the phenotypes were similar between pregnancies of nulliparous and multiparous women for both outcomes, with the largest difference observed for birth weight where there is a small shift in the distribution with smaller z-scores in first borns (Fig. S1). Nevertheless, we observed higher gestational duration variance in nulliparous compared to multiparous, and the opposite for birth weight (Table S1).

In the maternal nulliparity GWAS of gestational duration seven loci reached genome-wide significance, all with maternal only effects (Fig. 1, Fig. S2-S3, Table S2-S4); six of these loci have been previously identified (*WNT4, EBF1, TET3, ADCY5, HAND2* and *KCNAB1*) (6). The lead variant at the newly discovered locus is intronic to *CNTN6*, but this gene has no obvious relationship to gestational duration. In the multiparity GWAS, two loci reached the genome-wide significance threshold (located near *EBF1* and *KCNAB1*) (Fig. 1, Fig. S2-S3, Table S2-S4); both in high linkage disequilibrium (LD) (r^2^>0.85) with nulliparity variants in the same region. In the GWAS of the whole study population for the maternal genome on gestational duration, 11 loci reached genome-wide significance, two of these variants (*WT1* and *TET3*) had fetal only effects and one (*NDST4*) had maternal and fetal effects with opposite direction (Fig. S2-S3, Table S2-S4). Four of these 11 loci had never been associated with gestational duration before (r^2^<0.1, variants near *NDST4, WWTR1, HMGA1* and *WT1*) and all other loci were either previously identified or identified in the parity-stratified GWASs. Only the locus at *CNTN6* was significant only in the nulliparity GWAS and not in the whole study population GWAS. We highlight the lead variant at *WWTR1* (rs73155026, beta = −0.37 days, p-value = 1.6×10^−8^, MAF = 36.6%), which is fine-mapped as a splice quantitative Trait Locus (QTL) and expression QTL for WWTR1 in multiple tissues, supporting its role as the causal gene. Loss of WWTR1 messenger ribonucleic acid (mRNA) expression is observed in cytotrophoblasts from placentas of extremely preterm newborns (22).

**Fig. 1.**
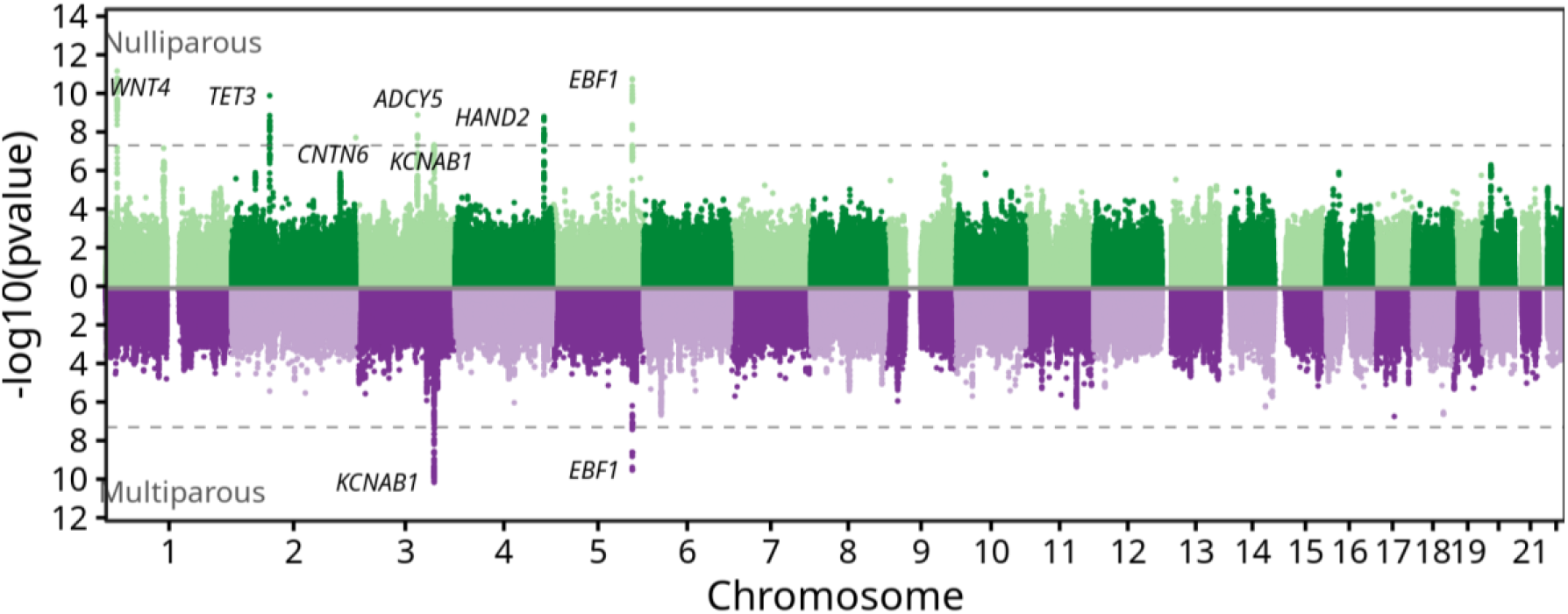
Miami plot illustrating the parity-stratified maternal GWASs on gestational duration. The top figure represents the GWAS in nulliparous women (n= 24,284), while the bottom figure illustrates the GWAS in multiparous women (n=31,202). The x-axis shows the chromosome position and y-axis the two-sided p-value of the GWAS. The dashed line represents the genome-wide significance threshold (p-value = 5×10−8). Variants reaching the genome-wide significance threshold have the nearest protein-coding gene displayed.

For the fetal nulliparity GWAS of gestational duration (Fig. S4-S5, Table S3-S5), we identified one genome-wide significant locus (*ADCY5*), with both fetal and maternal effects in the same direction, and none in the multiparity GWAS. To note, the locus is intronic for *ADCY5*, which harbours maternal and fetal effects on both gestational duration and birth weight and fetal effects on placental weight, adding another layer of complexity in this region. A previous GWAS has shown that the locus associated with gestational duration colocalizes with *ADCY5* gene expression in the uterus and affects fat-mass-related traits in the maternal genome (6). For birth weight, the locus affects glucose-related traits in the fetal genome. The fetal GWAS of gestational duration in the whole sample identified three independent loci (only fetal effects, *IL1A, WT1* and *LSM4*; Fig. S4-S5, Table S3-S5, Note S1).

For birth weight, the number of genome-wide significant loci was larger in the multiparous GWAS for the maternal genome (3 vs 8, Fig. S6, Table S3, S6), while no difference was found in the fetal genome (14 vs 14, Fig. S7, Table S3, S7). In the maternal GWASs, four loci had a fetal only effect (*LEKR1, PTCH1, ESR1* and *HMGA1*) two loci a maternal and fetal effect in the same direction (*TCF7L2* and *CRYZL2P-SEC16B*), and one locus a maternal and fetal effect in the opposite direction (*MTNR1B*, Fig. S8, Table S4).

While in the fetal GWASs, four variants had a maternal and fetal effect in the same direction (*ESR1, CKS1B, HLA-B* and *SLC25A37*), and an additional four variants had a maternal and fetal effect in the opposite direction (*ADCY5, KIF11, TENT4B* and *HHEX*, Fig. S9, Table S4). All maternal loci (including those from the whole sample GWAS) had been previously identified in (2), except for a signal at the *HLA-B* gene region associated both in multiparous women and in the GWAS using the whole sample, and a signal at the *CRYZL2P-SEC16B* region associated in the GWAS using the whole sample (Fig. S6 and Table S6). With regards to the fetal GWAS of birth weight, we discovered two associated loci in nulliparous women (*NEDD4* and *LEO1*) and two loci in multiparous woman (*METTL11B* and *CDC73*) which has not been identified previously (Fig. S7 and Table S7). In the whole sample GWAS we identified a total of 33 loci, seven of which were newly discovered (*NEDD4, SLC2A1, SIK2, HLA-B, SLC35D1, MSI2*, and *EDEM3*). As part of the identified novel loci, three lead SNPs were in strong LD with missense variants affecting *NEDD4* (rs8033275, beta = −0.05 z-score, p-value = 1.4×10^−10^, MAF = 22.5%), *WDR78* (rs2755253, beta = 0.03 z-score, p-value = 9.2×10^−9^, MAF = 32.7%) and *EDEM3* (rs78444298, beta = −0.10 z-score, p-value = 4.7×10^−8^, MAF = 2.0%). *NEDD4* regulates IGF-1 signalling and controls mice growth during development (23).

Using data from the Avon Longitudinal Study of Parents and Children (ALSPAC, n = 6,014-6,721 for whole study population across outcomes, Table S8) we were able to replicate the direction of the effect of 103 of 110 testable associations, with the exceptions: *CNTN6* for gestational duration in the maternal genome; *ESR1* and *CRYZL2P-SEC16B* for birth weight in the maternal genome; and *CDC73, YKT6, IL17REL* and *EDEM3* for birth weight on the fetal genome (Table S9). A sign test indicated that the number of variants with the same direction of effect were greater than expected under the null hypothesis (binomial test, p < 2.2×10^−16^, proportion same direction = 0.95, 95% confidence interval (CI): 0.87–0.97 null hypothesis: observations would match the direction by chance - 0.5).

Because nulliparous and multiparous mothers differ in more factors than parity itself, we also compared the maternal genetic effects of the identified variants in mothers with the first two pregnancies available in MoBa (n = 5,895 for gestational duration and n= 6,942 for birth weight) and after accounting for maternal age. For gestational duration, out of the 12 lead SNPs identified, 9 had a larger absolute effect in the first pregnancy (Fig. S10). With a lenient Bonferroni correction (0.05/12), three loci reached significance threshold in the first pregnancy (*TET3, KCNAB1* and *NDST4*), whereas no variants were significant in the second pregnancy. For birth weight 9 out of 14 loci had a larger absolute effect in multiparous woman (Fig. S10). With a lenient Bonferroni correction (0.05/14), two locus reached significance threshold in the first pregnancy (*ATXN2* and *YKT6*), whereas six variants were significant in the second pregnancy (*LMO1, GLIS3, LEKR1, SLC38A4, HMGA1*, and *YKT6*). Overall, we see the same trend as in the main study population, suggesting that factors specific to nulliparous/multiparous women are not the drivers of the differences in effect sizes.

The correlation between the effects of the genome-wide significant variants from the parity-stratified GWASs were lowest for gestational duration using the maternal genome (not assessed for fetal genetic effects on gestational duration due to a small number of significant loci). We did not observe variants with opposite effect direction when splitting by parity, suggesting, if anything, that nulliparity affects the magnitude of effect (Fig. S11). However, we note that it is also possible that the variants have an effect in pregnancies of nulliparous women and a null effect in pregnancies of multiparous women, or vice-versa (a distinction that cannot be made based solely on the correlation between effect sizes).

To ensure that the analyses were not influenced by factors related to fertility or selection bias, we conducted GWASs of participating with the first or later pregnancies (case-control study) for the maternal and fetal genome. No variants reached genome-wide significance (Fig. S12), indicating no gross genetic differences between first and later-born.

We also investigated whether the gestational duration (in days) or birth weight (in z-score) in previous pregnancies influenced the estimated effects of the genetic variants we identified. Using mothers with their two first deliveries registered in MoBa (n = 3,021-6,942 mothers), the genetic effects for the second pregnancy with and without adjusting for the gestational duration or birth weight of the first pregnancy were overall consistent (r = 0.92-1, Fig. S13).

Overall, the parity-stratified GWASs highlighted shared and distinct loci for both gestational duration and birth weight, with no clear pattern across outcomes. However, genetic variants reaching genome-wide significance had the same effect direction in the nulliparity and multiparity GWASs.

### Two genetic variants previously associated with gestational duration interact with nulliparity

We then focused on the gene × nulliparity interactions for genetic variants identified in previous studies (2–6) (n_snps_ = 3 - 189) due to the comparatively lower sample size of our cohort and the multiple hypothesis-testing burden. Based on Bonferroni correction adjusting for the total number of variants analysed for each phenotype and genome, two genetic variants for gestational duration reached the significance threshold for gene × nulliparity interactions (Fig. 2, Fig. S14, Table S10, and p-value thresholds in Table S11). Specifically, the maternal lead variants for gestational duration in *ADCY5* and *WNT4* had a larger absolute effect size in nulliparous compared to multiparous women. The interaction between the genetic variant and nulliparity remains unchanged when also adjusting for the term gene × maternal age (given the one more degree of freedom) (Table S12).

**Fig. 2.**
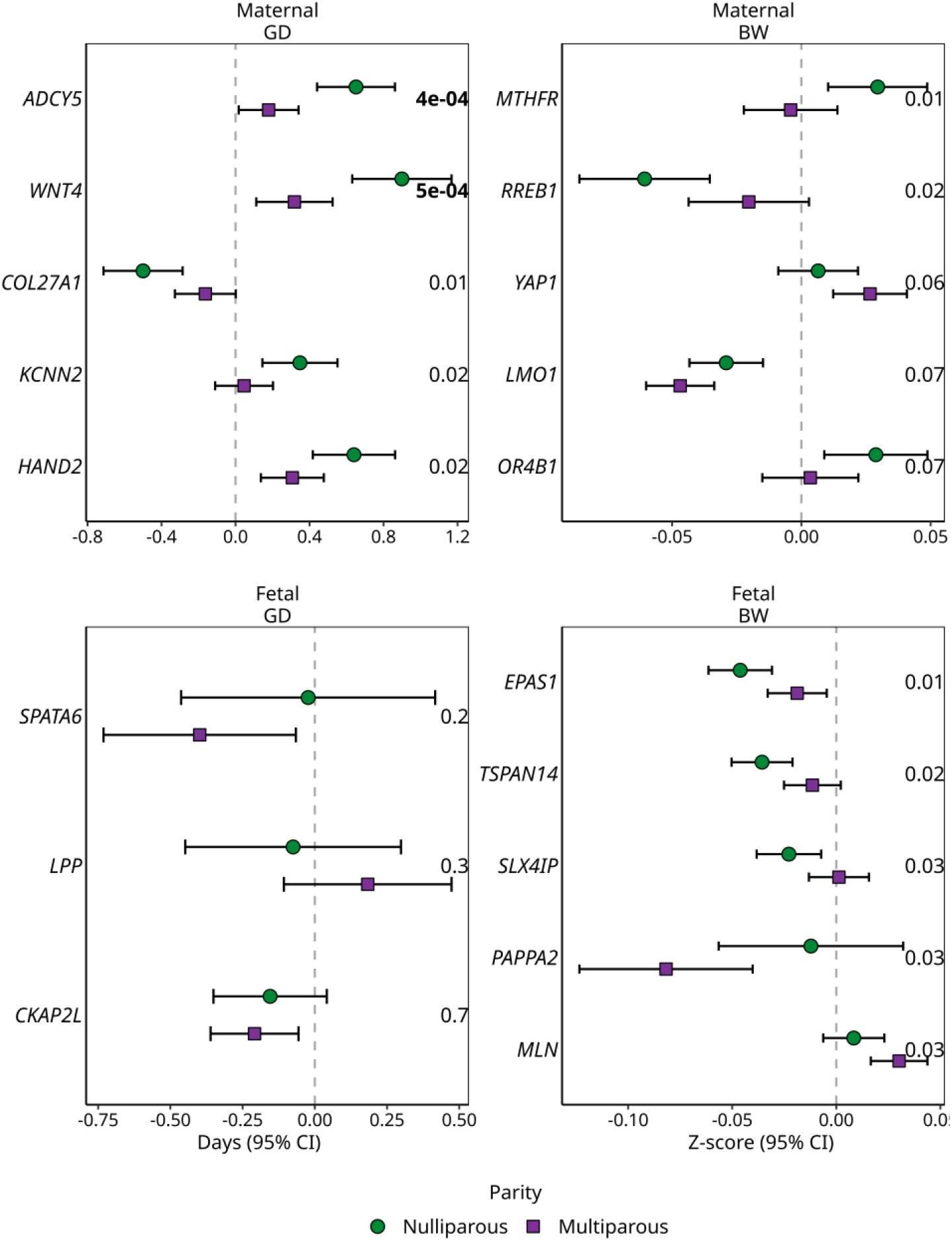
Parity-stratified effect sizes (n ~ 28,000 mothers/ children) for the top five variants with lowest interaction p-value (SNP × parity) for gestational duration (GD) and birth weight (BW) in the maternal and fetal genomes. The green circles are the effect sizes in pregnancies of nulliparous woman and the purple square are the effect sizes in pregnancies of multiparous woman. The error bars are the 95% confidence interval. Variants are displayed as the nearest protein-coding gene on the y-axis. The interaction p-value for each variant is displayed to the right. Only variants previously associated with the specific outcome in (2–6) were included (nsnps = 3 - 189). For maternal gestational duration, maternal birth weight and fetal birth weight the remaining variants are illustrated in Fig. S14. Two genetic variants reached the Bonferroni-corrected significant threshold, with their p-values highlighted in bold (for p-value thresholds see Table S11).

*WNT4* is important for the development of the female reproductive system (24–26) and breast cancer (27). In mouse mammary cells, increasing parity decreased *WNT* signalling partly due to a more than threefold reduction in the *WNT4* ligand expression (28). This reduction of *WNT4* expression was believed to be linked to a decrease in the number of oestrogen/progesterone receptors in luminal cells in multiparous mice. Moreover, previous functional analysis of the *WNT4* locus associated with gestational duration indicated that the likely causative variant alters the binding of the *ESR1* oestrogen receptor by an improved binding capacity for the minor allele (29). If there is also a decrease in the number of oestrogen/progesterone receptors in uterus cells in multiparous women, it is possible that this variant plays a less relevant role in multiparous compared to the nulliparous women.

### The cumulative effect of gestational duration-associated variants is modified by nulliparity

We first assessed whether the aggregated effect of previously identified variants (2–6) was modified by nulliparity using random forest models. We created random forest genetic scores to avoid splitting the data into training and testing sets, which would reduce the sample size available for testing the interaction between genetic scores and nulliparity, and allow for gene-gene interactions.

Among all the phenotype-genome combinations, only the random forest genetic scores of the maternal genome on gestational duration showed a significant interaction with nulliparity (p-value interaction=5×10^−5^, Table S13). The interaction indicated that the effect of the random forest genetic score on gestational duration is larger in nulliparous compared to multiparous women (Fig. 3A, Table S13). Since the random forest genetic score only included genetic variants as features, this interaction indicates that known variants associated with gestational duration have a significantly higher genetic effect on gestational duration in nulliparous women. To evaluate whether the identified interaction originates primarily from the maternal or fetal genome, the maternal genetic variants were examined by splitting these into maternal non-transmitted and paternal transmitted in separate random forest models. The interaction was strongest and the prediction accuracy highest, in the model including the maternal non-transmitted (MnT) alleles using mother-child duos in the whole study population (Fig. S15, r^2^ MnT: median 0.0027 [2.5th percentile 0.0026, 97.5th percentile 0.0029] and r^2^ paternal transmitted (PT): median - 3.0×10^−5^ [2.5th percentile −4.3×10^−5^, 97.5th percentile −1.8×10^−5^]), indicating a primary maternal effect. Furthermore, in the random forest models derived from the whole population that included both genetic and environmental features the interaction for gestational duration in the maternal genome remained significant (Table S14). This result was supported when splitting the random forests by parity, which had a higher predictive accuracy (r^2^) in nulliparous women for the maternal genome on gestational duration (the 95% credible intervals did not overlap). We also observed higher predictive accuracy for the fetal genome random forest genetic scores on birth weight in nulliparous compared to multiparous women (Fig. 3B, Table S15). Conversely, for gestational duration in the fetal genome we observed higher predictive accuracy in pregnancies of multiparous woman.

**Fig. 3.**
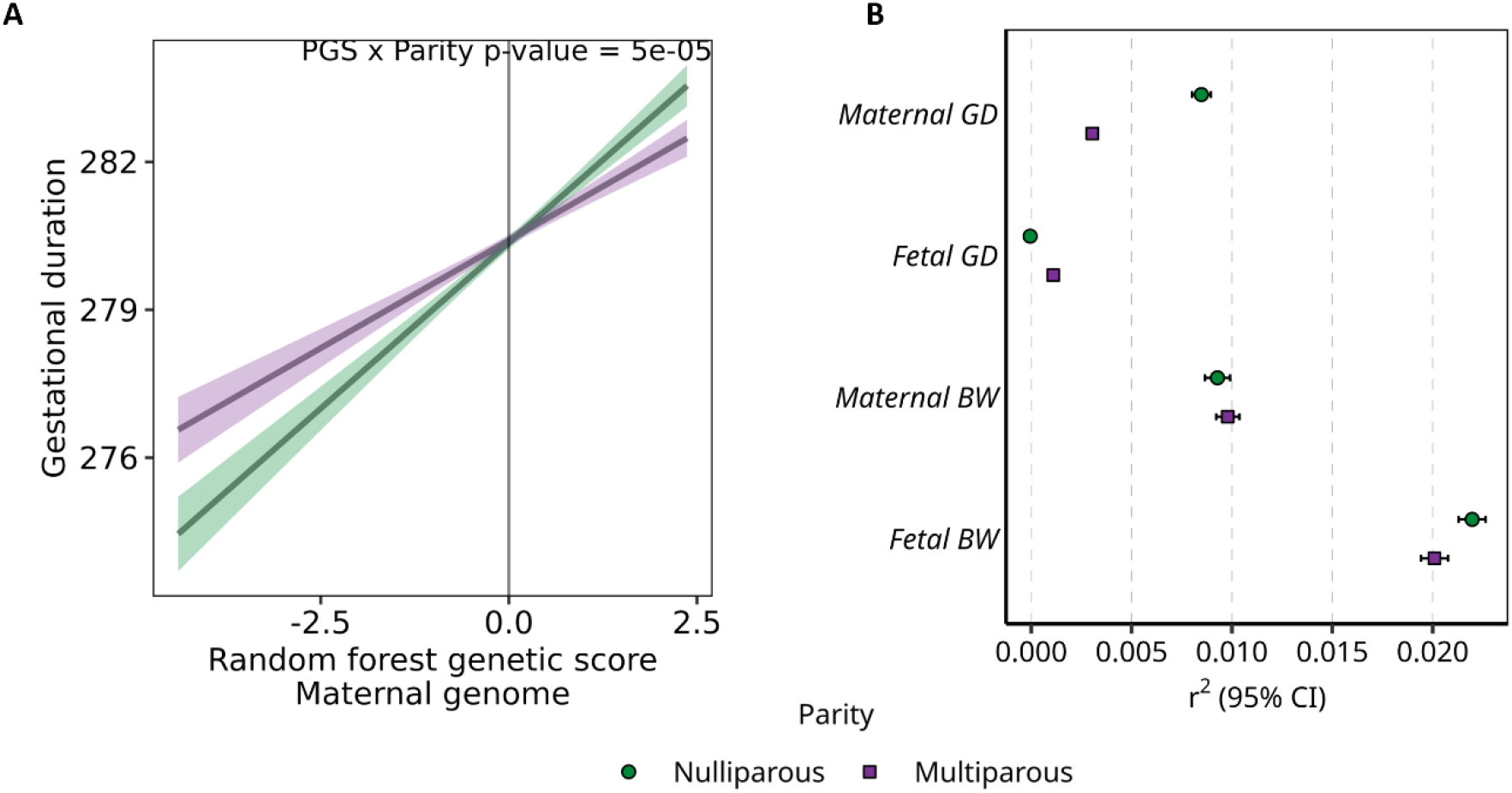
The interaction between the maternal random forest genetic score and nulliparity on gestational duration, along with the prediction accuracy of each phenotype and genome in random forest models stratified by parity. (A) The interaction between the maternal random forest genetic score and nulliparity on gestational duration. The slopes highlight the effect of the random forest genetic score on gestational duration by parity (green line: nulliparity (n=24,284), purple line: multiparity (n=31,202)) and are based on the estimates from the linear model used when testing the interaction (Table S13). The x-axis shows the maternal random forest genetic score and the y-axis the gestational duration. The interaction p-value is displayed in the top right of the figure. (B) Prediction accuracy (r2) of each phenotype (gestational duration = GD, birth weight = BW) and genome in random forest models stratified by parity (n~28,000 mothers/ children) and constructed using hard-coded genotypes of variants reaching genome-wide significance in (2–6). Each point represents the prediction accuracy from one random forest model. The green circle is the prediction accuracy in pregnancies of nulliparous woman and the purple square are the prediction accuracy in pregnancies of multiparous woman. The 95 % credible interval was generated from 10,000 of r2 on out-of-bag predictions for the tuned hyperparameters in each model.

For birth weight on the maternal genome, the 95% credible intervals overlap between nulliparous and multiparous women. Notably, the largest fold change in prediction accuracy between nulliparous and multiparous woman was observed for gestational duration in the maternal genome, with almost a 3-fold increased predictive accuracy (Table S15). Overall, the predictive accuracy of the random forest genetic scores was low for all outcomes, except for the fetal genome on birth weight. Adding known environmental factors as part of the random forest predictions increased the variance explained for all phenotypes except for gestational duration using the fetal genome in nulliparous women (Note S2).

To assess whether the stronger genetic effect on gestational duration we have observed in nulliparous women is not limited to previously identified variants, we examined interactions between a PGS of gestational duration with nulliparity in the ALSPAC cohort (n=6,281). Here, we constructed a genome-wide PGS of gestational duration including 1,120,697 variants, with weights derived from the current GWAS in MoBa (all women, nulliparous and multiparous). We observed a significant interaction between the PGS of gestational duration derived from the GWAS in all women and nulliparity on gestational duration in ALSPAC (p-value=7.06×10^−4^), with stronger effects in nulliparous compared to multiparous women (Fig. S16, Table S13) and after accounting for the correlation between maternal and fetal genotype. This result validates the abovementioned random forest scores at the genome-wide level in an independent cohort.

We then examined the associations between the PGS of gestational duration derived from nulliparous and multiparous women on gestational duration in nulliparous and multiparous women from ALSPAC, per separate (n=2,855 and 3,468, respectively). Overall, we observe that the PGS of gestational duration from multiparous women performs poorly both in nulliparous and multiparous women (R2=0.0071 and R2=0.0071 respectively, Table S16). The PGS of gestational duration from nulliparous women, however, explains 0.0127 and 0.0081 of the variance of gestational duration in nulliparous and multiparous women, respectively.

Overall, we observe stronger genetic effects and improved predictive accuracy of gestational duration in nulliparous women using genetically predicted gestational duration using the maternal genome.

### Parity-dependent effect sizes and differences in the total genetic variance by parity

Next, we estimated genetic correlations and heritability for each phenotype and between pregnancies of nulliparous and multiparous woman using LD score regression (30, 31). The genetic correlation between the GWAS in nulliparous and multiparous women for any phenotype using either the maternal or fetal genome ranged from 0.66–0.75 (Fig. 4).

**Fig. 4.**
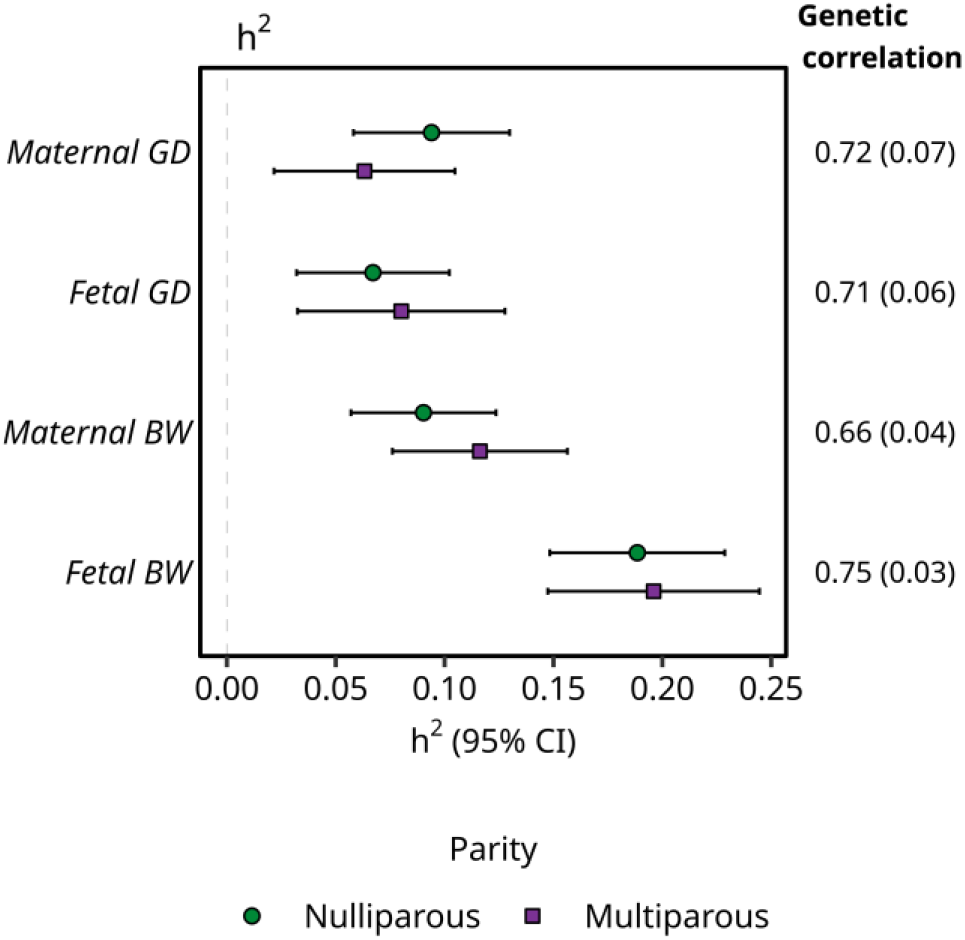
SNP heritability for gestational duration (GD) and birth weight (BW) in the maternal and fetal genomes by nulliparity (n~28,000 mothers/children). The green circles are the heritability estimates in nulliparous women and the purple square are the heritability estimates in multiparous women. The error bars are the 95% confidence interval. On the right side of the figure, the genetic correlation between nulliparous and multiparous women is displayed with its standard error in parentheses.

Compared to the correlations reported in (32), which analysed 33 traits and 10 environmental variables for gene × environment interactions (with genetic correlations ranging from 0.85 to 0.98), the correlations in this study are relatively low, implying parity-specific genetic effects. Furthermore, the imperfect genetic correlations are in concordance with the parity-stratified GWASs, which suggested different lead SNPs by nulliparity (Fig. 1, Fig. S2, S4, S6, S7).

Narrow-sense heritability estimates were not significantly different between nulliparous and multiparous women for any of the two phenotypes using the maternal or fetal genomes (Fig. 4, Table S17). However, the number of discovered loci for both gestational duration and birth weight do vary by parity, except for the fetal genome. Given the low deviation in sample size by parity (6,370–8,272 more individuals in multiparous), we speculate that the difference in discovered loci by parity arises from the underlying genetic variance in each parity group. This can be further supported by comparing the number of variants reaching the significance threshold; we observe a higher proportion of associated loci per study participants in the parity group with more significant variants in the maternal genome for both gestational duration and birth weight compared to previous studies with larger sample sizes (Table S18).

## Discussion

In previous GWASs of gestational duration and birth weight whether women had at least one previous pregnancy or not was not considered. In this study, we investigated whether nulliparity modifies the maternal or fetal genetic effects on these pregnancy outcomes. Our findings show that the identified loci and their numbers were moderately different when stratifying the GWASs by parity. This finding is supported by maternal genetic scores explaining higher gestational duration variance in nulliparous women in an independent sample.

We observed the strongest indication of effect modification by parity for the maternal genome on gestational duration. Our findings suggest that the magnitude of genetic effects is larger in nulliparous woman, and that this effect is not limited to previously identified variants. However, the molecular mechanisms regulating the timing of delivery in humans are poorly understood, complicating the biological interpretation of these results. During a woman’s first pregnancy and delivery, numerous physiological first-time changes occur in the body, this could possibly explain the larger genetic effects for gestational duration in the maternal genome. One example is the growth and expansion of the uterus. Parity is associated with smaller uterine size in nulliparous women (12, 33). After the first pregnancy, the muscle fibres become stretched and will more easily expand for further pregnancies, which can result in the genetic control to expand the uterus being less important in subsequent pregnancies. This is supported by the variant near *WNT4*, which in our study showed a larger effect in nulliparous women and has been correlated with an increase in uterine size by 35% in mice (34). It is difficult to conclude anything about the fetal effects on gestational duration given the few loci known to be associated with this phenotype.

While our findings for gestational duration were robust, the picture for birth weight is blurrier. Even if we have identified larger number of loci in the GWAS of birth weight in both the maternal and fetal genomes compared to gestational duration, we find less consistent modification effects by nulliparity. However, this may have arisen as a bias induced by the fact that data from previous GWAS on birth weight were largely based on data from the UK Biobank, where birth weight data is from nulliparous women. Moreover, we know that there is an interplay between the two studied phenotypes, where gestational duration is a key determinant of birth weight. As previously discussed, we hypothesise that the genetic control to expand the uterus does not need to be as large in multiparous women since the uterus expands more easily after the first pregnancy. Animal studies have shown that uterine capacity influences birth weight; for instance, large breed sheeps were lighter and smaller when born to small breed dams (35). Easier expansion of the uterus in pregnancies of multiparous women may also result in greater flexibility for the fetus to adjust its growth. This could be one reason for larger mean birthweight in multiparous women, and maybe larger genetic effects in this subgroup for birth weight (although we obtained limited evidence for this speculation). Recent work has examined the association between a maternal genetic score of birth weight and estimated fetal growth in nulliparous and multiparous women separately (36). While the interactions between the maternal genetic score and nulliparity were not formally tested, the 95% CI of the effects sizes of the genetic score on fetal growth overlapped between nulliparous and multiparous, suggesting no strong interactions.

Our results highlight the importance of considering nulliparity in genetic studies of pregnancy outcomes. We suggest that nulliparity modifies the genetic variance of gestational duration. It is important to study a homogeneous population, and nulliparity should be considered in future genetic research related to pregnancy outcomes. For example, genetic studies focusing on gestational duration could prioritize nulliparous women to increase discovery per individual recruited. While we have replicated the effect modification of nulliparity in ALSPAC, additional replication in other cohorts that include individuals of non-European ancestries should be performed to strengthen the results shown in this paper. A strength of this study is the availability of genotype data from both mothers and offspring in >50,000 samples in a single cohort; however, the sample size is comparatively small to genetic studies of non-pregnancy phenotypes. Increasing sample size might reveal additional loci with different effects across pregnancies and might help resolve parity effects that in this study were difficult to interpret, particularly in relation to birth weight.

To conclude, we show that nulliparity modifies the maternal genetic effect on gestational duration, with stronger predictive accuracy and additional genetic loci discovered in nulliparous compared to multiparous. Our study highlights the complexity of the genetics of pregnancy duration and other pregnancy outcomes, where not only two genomes coexist in a single environment, but also the number of previous pregnancies might modify the effect of these genomes.

## Materials and Methods

### Study cohort

MoBa is a population-based pregnancy cohort conducted by the Norwegian Institute of Public Health. Participants were recruited from all over Norway from 1999 to 2008. The women consented to participation in 41% of the pregnancies. The cohort includes approximately 114,000 children, 95,000 mothers and 75,200 fathers (37, 38). Blood samples were obtained from both parents during pregnancy and from mothers and children (umbilical cord) at birth (39). MoBa participants responded to questionnaires during and after the pregnancy and have been linked to Norwegian registries. The current study is based on version 12 of the quality-assured data files released for research in 2019. The MoBa data used in this study was linked to data from the Medical Birth Registry of Norway (MBRN; a national health registry containing information about all births in Norway from 1967 onwards) (40). In this study, genotyping data from 207,569 unique individuals of European ancestry (76,577 children, 53,358 fathers, and 77,634 mothers) and 6,981,748 SNPs that passed the MoBaPsychGen pipeline were analysed (41). Genotyping, pre-imputation quality control (QC), phasing, imputation, and post-imputation QC have previously been described in full (41).

### Sample selection

To be included in this study participants needed to be registered in both MoBa and MBRN. For mothers with several pregnancies included in this set of genotyped MoBa samples (n=9,923), one random pregnancy per mother was included when analysing the maternal genome to ensure independent samples for the analysis. This is important for the parity-stratified analyses as it prevents any single mother from being represented in both parity groups. For the analysis using the maternal or fetal genome, individuals were restricted based on relatedness; not allowed to be full siblings (king-cutoff 0.1327) (42). For the two phenotypes, the analysis was restricted to singleton live births conceived without the use of assisted reproductive technology. Records were excluded if the sex assigned at birth of the child was unknown, the child was born with malformations, or if there was an early perinatal death. For definitions and specific criteria within each phenotype and the exposure parity see below.

#### Gestational duration

Gestational duration is based on the records from MBRN, it is measured in days and based on ultrasound estimation in the mid second trimester. However, if ultrasound is not available the gestational length is calculated from the last menstrual period (in this cohort more than 98% of the pregnancies were estimated with ultrasound). The analysis was restricted to spontaneous labor, with a gestational duration ranging between 154 to 308 days. Children dying within one year from birth were not included.

#### Birth weight

The offsprings birth weight (in gram precision) was obtained from the MBRN. The analyses of the birth weight were limited to gestational durations between 259 and 300 days. Children with a birth weight within ± 5 standard deviations from the mean birth weight were included. The birth weight was transformed to z-scores for all analyses using the mean and standard deviation from the study population.

#### Parity and nulliparity

Parity is defined as the number of times a woman has previously given birth to a child, live or stillborn. In MBRN parity is determined as the greater of “Previous births reported by the mother” and “Previous births recorded in the MBRN”. Previous births were registered in MBRN from week 16 (112 days) between 1967 and 2001, and from 2002 onwards, they were registered from week 12 (84 days). The counting of parity is zero-based, meaning that the first pregnancy is parity zero (nulliparous) and so on. Mothers with uncertain parity were removed to avoid potential errors in parity: mothers who gave birth to more than one child with the same parity, and cases where the parity did not correspond to the order of delivery (for example, a parity zero occurred after a parity one). Throughout the paper, parity will be defined as a dichotomous trait by nulliparity: pregnancies of nulliparous vs. multiparous women.

### Replication cohort: ALSPAC

We attempted to replicate the loci reaching the genome-wide significant threshold in any of the 12 GWASs (Table 1) using the ALSPAC cohort. ALSPAC is a longitudinal birth cohort, which has been described in detail elsewhere (43, 44). In short, pregnant women resident in Avon, United Kingdom with expected dates of delivery between 1st April 1991 and 31st December 1992 were invited to take part in the study. 20,248 pregnancies have been identified as being eligible and the initial number of pregnancies enrolled was 14,541. Of the initial pregnancies, there was a total of 14,676 fetuses, resulting in 14,062 live births and 13,988 children who were alive at 1 year of age. Of the original 14,541 initial pregnancies, 338 were from a woman who had already enrolled with a previous pregnancy, meaning 14,203 unique mothers were initially enrolled in the study. Pregnancy information was retrieved from questionnaires (generation G0, mothers), the cohort profile and obstetric data. Please note that the study website contains details of all the data that is available through a fully searchable data dic-tionary and variable search tool (http://www.bristol.ac.uk/alspac/researchers/our-data/).

**Table 1.** Sample sizes of the 12 genome-wide association studies preformed.

| Phenotype | Genome | n whole study population | n nulliparous woman | n multiparous woman |
| --- | --- | --- | --- | --- |
| Gestational duration | Maternal | 55,486 | 24,284 | 31,202 |
|  | Fetal | 50,728 | 22,184 | 28,544 |
| Birth weight | Maternal | 64,330 | 28,029 | 36,301 |
|  | Fetal | 58,894 | 25,648 | 33,246 |

Mothers (G0) were genotyped using the Illumina Human660W-quad array while the off-springs (G1) were genotyped using Illumina HumanHap550 quad chip platforms. Imputation procedures for ALSPAC have been described in detail in the ALSPAC omics data catalogue. From this data set, 6,014-6721 mothers or offsprings passed genotype QC and inclu-sion/exclusion criteria for the whole study population (nulliparous + multiparous) across outcomes and were included in the replication analysis.

Analyses were restricted to singleton pregnancies with live-born infants surviving to one year of age. We excluded pregnancies with stillbirth, known fetal complications, or conception via in vitro fertilisation, as well as pregnancies delivered following induction of labor or caesarean section.

### Genome-wide association studies

The GWASs were performed using REGENIE (v3.2.5) (45) with analyses consisting of three groups: whole sample, pregnancies of nulliparous women, and pregnancies of multiparous women for the two phenotypes (gestational duration and birth weight) when analysing either the maternal or fetal genome. This resulted in a total of 12 GWASs (Table 1). For gestational duration, the regression models were adjusted for maternal age, fetal sex assigned at birth, genotyping chip and the ten first genotype principal components. For birth weight the GWASs were adjusted for the same factors, in addition to gestational duration. Top lead genetic variants were those with an association p-value below the genome-wide significance threshold (5×10^−8^) with a radius of 1.5×10^6^ base pairs.

### Haplotype analysis to resolve effect origin

We resolved whether the variants reaching the genome-wide significant threshold were of maternal, fetal, or maternal and fetal origin using mother-child duos and phased genotype data (estimated haplotypes) as previously described (46). In short, we used SHAPEIT5 (47) to estimate haplotypes from hard coded genotypes using the --pedigree option with a pedigree from our parent-offspring duos or trios. This tool uses a hidden Markov model in combination with information from genetic relatives to refine phase calls and assign parents to haplotypes. From the phased offspring data, we obtained maternal transmitted (MT), MnT and PT alleles for each of the lead SNPs. For each of these, we fit the following linear regression model (Eq. 1):

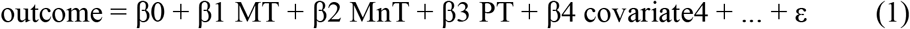

A variant was classified as having a maternal-only effect when PT 95% CI overlapped zero and the MnT 95% CI did not overlap zero. A fetal-only effect was defined when MnT 95% CI overlapped zero and the PT 95% CI did not overlap zero, or when only MT 95% CI did not overlap zero. Variants were classified as having both maternal and fetal effects when the 95% CI for both PT and MnT did not overlap zero. These models were run using data from 17,869 up to 50,479 mother-child duos across outcomes, genotypes, and parity groups (Fig. S3, S5, S8, S9).

### Replication analyses of identified loci

Given the differences in sample size between MoBa and ALSPAC, replication of the associations between the individual genetic variants and the phenotypes was only attempted in terms of effect direction. Association analyses were done under an additive genetic model using REGENIE v4.1 including the same covariates for each outcome as for the discovery population.

### Sensitivity analysis of participating with the first or later pregnancies

To ensure that the analyses were not influenced by factors related to fertility or selection bias, we conducted GWASs of participating with the first or later pregnancies (case-control study, nulliparity as outcome) for the maternal and fetal genome using REGENIE (v3.2.5). The regression models were adjusted for maternal age, genotyping chip and the ten first genotype principal components.

### Sensitivity analyses using the same women with information from first and second pregnancies

We conducted further sensitivity analyses to assess whether maternal genetic effects on gestational duration were also stronger in the first and second pregnancies (and the opposite for birth weight) limiting the analyses to mothers for which we had information from the two pregnancies. This analysis was performed because nulliparous and multiparous women are known to differ in other factors different to parity. For this, we used data from MoBa from 5,895 women from whom we had the duration of pregnancy from the first and subsequent pregnancies. In this subset of women, we estimated the effects of the identified genetic variants on gestational duration in the first and second pregnancy per separate.

One of the strongest predictors of a second pregnancy’s outcomes is the outcome from the first pregnancy. To this end, we used the same set of women as above (women from MoBa for whom we had data from the first and second pregnancies) and evaluated whether adjusting for first pregnancy outcomes modified the association between the identified maternal and fetal genetic variants and gestational duration and birth weight, after adjusting for the same covariates as in the main GWAS. An example model looks as follows (Eq. 2):

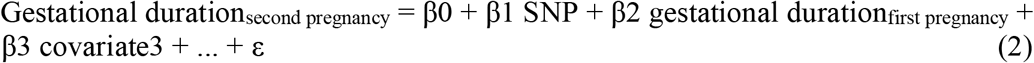

### Gene × environment interaction analysis for previously identified loci

We assessed whether previously discovered loci associated with gestational duration or birth weight using either the maternal or fetal genome for gene × environment interactions on the whole sample for each phenotype and genome was conducted with REGENIE (v3.2.5). We used a linear regression model that included nulliparity, SNP and the interaction SNP × nulliparity terms, adjusting for the same covariates as in the GWASs explained above.

As a sensitivity analysis, we further adjusted for a SNP × maternal age term to capture potential confounding in the interaction SNP × nulliparity using the same samples and model as above mentioned.

### SNP proxies for missing variants

Some of the previously identified variants associated with gestational duration or birth weight did not pass quality control or were not part of the imputation panel. To retain the maximum information, variants present in our genotype data within +/-550 000 base pairs of the missing top SNP were extracted. The LD pattern of each genomic region was then explored using the LDassoc Tool from LDlink (48). If a variant within the genomic region had an r^2^ > 0.3 to the missing top SNP, this SNP was used as a proxy. When several variants reached an r^2^ > 0.3 with the missing variant, the variant with the largest r^2^ was selected. After searching for proxies there were still some top SNP missing (Table 2).

**Table 2.** Genetic variants previously associated with the phenotypes in this study’s genotype data.

| Phenotype | Genome | Sample size in references | Top SNPs still missing | Number of Top SNPs left |
| --- | --- | --- | --- | --- |
| Gestational duration | Maternal | (5): n = 68,732-98,370<br>(6): n = 195,555 | 5* | 27 |
|  | Fetal | (3): n = 21,262<br>(4): n = 84,689 | 1 | 3 |
| Birth weight | Maternal | (2): n = 230,069 | 4 | 74 |
|  | Fetal | (2): n = 321,223 | 10† | 189 |

\* Note that for the maternal genome for gestational duration two of the missing SNPs are located on the X chromosome, which is not included in the release we had of MoBaPsychGen.
† Note that for the fetal genome for birth weight three of the missing SNPs are located on the X chromosome, which is not included in the release we had of MoBaPsychGen.

### Expression QTL loop-ups

To connect lead SNPs to potential causal genes, we looked up each of them in the eQTL Catalogue (49). This resource provides a uniformly processed set of expression and splice QTLs in 127 datasets from a multitude of tissues and cell types. More specifically, we searched whether the lead SNPs we have identified were part of fine-mapped sets, and if so, which were the potential target genes and in which tissues/ cell types.

### Polygenic prediction of pregnancy outcomes using random forest models

The goal of the random forest genetic score was to reflect the genetic effects on the two phenotypes based on genetic variants reaching genome-wide significance in previous meta-analyses (2–6) (n up to 321,223 across studies). To prevent splitting the data into entirely separate training and testing sets, which would reduce the sample size for testing the gene × nulliparity interaction, out-of-bag predictions were applied on the random forest models (50). This approach allowed us to use the full data both when constructing the genetic prediction and when assessing if nulliparity interacts with the genetic prediction (top variants across the entire genome). Moreover, a benefit of using random forest is that it allows to capture non-linear associations.

Random forest models were created to predict the two phenotypes using maternal or fetal hard-coded genotypes (coded as 0, 1, 2) of the genetic variants previously reaching genome-wide significance in the GWASs (2–6) for each outcome. Each phenotype (gestational duration and birth weight z-score) was residualized as the mean plus the individuals’ residual from a linear model adjusting for the same covariates as in the GWASs mentioned above: maternal age (for gestational duration model), gestational duration (for birth weight model), fetal sex assigned at birth, genotyping chip and the ten first genotype principal components (for both). The prediction accuracy of the random forest models represents the genetic predictability of the phenotype and was estimated as 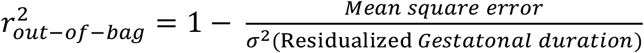.

From the random forest models, we obtained genetically predicted residualized gestational durations/birth weight z-scores, we will refer to these as random forest genetic scores. Since random forests have a random component (each decision tree in the forest is created using a random subset of the dataset and each node in a tree is created using a random subset of variables), a mean random forest genetic score was created for each individual based on 10 000 out-of-bag predictions with the tuned hyperparameters. The mean random forest genetic score was used to test for gene × nulliparity interaction on each outcome in a linear model for the same samples used to create the random forest model (Phenotype = β0 + β1 random forest genetic score + β2 nulliparity + β3 random forest genetic score * nulliparity, the random forest genetic score was centered to have a mean of zero).

Furthermore, two similar random forest models of the residualized phenotypes were constructed separately for nulliparous and multiparous women to estimate parity-specific genetic predictability for each outcome. We also used the whole sample and split by nulliparous and multiparous women, to construct random forest models not only using genetic features but also including the abovementioned covariates (i.e., the outcome was non-residualized). All models created for each phenotype are summarized in Table 3.

**Table 3.** Description of random forest models (per phenotype-genome combination)

| Model description |  |  |
| --- | --- | --- |
| Population | Features | Outcome |
| Whole study population | Genetics only | Residualised |
| Nulliparous woman | Genetics only | Residualised |
| Multiparous woman | Genetics only | Residualised |
| Whole study population | Genetics and environmental | Non-residualised |
| Nulliparous woman | Genetics and environmental | Non-residualised |
| Multiparous woman | Genetics and environmental | Non-residualised |

Models were built in R using the ranger package (51). For the performance of the random forest, hyperparameters were tuned (Supplementary Method). To estimate how well each model performs in terms of genetic predictability, a 95 % credible interval was generated from 10 000 r^2^ on out-of-bag predictions from each model, using the tuned hyperparameters.

We further tested the effect origin for models showing a significant interaction between the random forest genetic score and nulliparity on the residualised outcome. We modeled haplotype-based genetic scores to assess if the effect originated from the maternal or fetal genome, using random forest models in the whole study population. The transmitted and non-transmitted allele from mother-child duos (coded as 0, 1) were used in separate models, which were consistent with the main models described above.

### Maternal polygenic scores of gestational duration and validation in ALSPAC

We further constructed a PGS of gestational duration using PRS-CS v1.1.0 (52). It is a Bayesian polygenic prediction method that estimates posterior SNP effect sizes under a continuous shrinkage prior while accounting for LD using an external reference panel. We used MoBa GWAS summary statistics on gestational duration derived from the maternal genome as the training dataset and generated posterior SNP weights in PRS-CS using an ancestry-matched LD reference panel (1000 Genomes European samples). We then computed PGS in target dataset – ALSPAC – by summing the imputed allele dosages weighted by the PRS-CS posterior effect estimates, after harmonizing alleles between summary statistics and the target genotypes and excluding ambiguous strand SNPs when necessary. In total, we computed three different PGS (all samples, nulliparous and multiparous GWAS). The PGS were z-standardized within ALSPAC prior to downstream association analyses. The PGS were used to assess PGS x nulliparity while also accounting for the correlation between maternal and fetal genotype (Eq. 3):

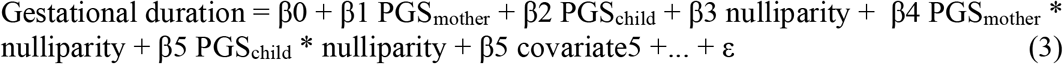

where the PGS_child_ is based on the maternal summary statistics. The PGS were also used to estimate the variance explained of the PGS in each subgroup of women (i.e., nulliparous and multiparous). We not only estimated the variance explained within each of the subgroups, but also across subgroups (i.e., the PGS derived from nulliparous was assessed in both nulliparous and multiparous women, and vice-versa).

### Heritability and genetic correlation

SNP heritability was estimated with LD score regression (LDSC) (v 1.0.1) (30), using pre-computed LD Scores from 1000 Genomes European data and GWAS summary statistics as input. The genetic correlation between nulliparous and multiparous women were also estimated using LDSC (31) for the two phenotypes and the maternal and fetal genome.

### Ethical approval

#### Norwegian Mother, Father and Child Cohort Study

Informed consent was obtained from the study participants. The study protocol was approved by the administrative board of the Norwegian Mother, Father and Child Cohort Study which is led by the Norwegian Institute of Public Health. MoBa is regulated by the Norwegian Health Registry Act. The establishment of MoBa and initial data collection was based on a license from the Norwegian Data Protection Agency and approval from The Regional Committee for Medical Research Ethics. The current study was approved by The Norwegian Regional Committee for Medical and Health Research Ethics (#2015/2425) and by the Swedish Ethical Review Authority (Dnr #2025-03908-02).

#### Avon Longitudinal Study of Parents and Children

Ethical approval for the study was obtained from the ALSPAC Ethics and Law Committee and the Local Research Ethics Committees. Informed consent for the use of all data collected was obtained from participants following the recommendations of the ALSPAC Ethics and Law Committee at the time. Participants can contact the study team at any time to retrospectively withdraw consent for their data to be used. Study participation is voluntary and during all data collection sweeps, information was provided on the intended use of data. Biological samples are collected in accordance with the Human Tissue Act (2004). The completion of a questionnaire, either on paper or online, was considered to be written consent from participants to use their data for research purposes. The study was additionally approved by the Swedish Ethical Review Authority (Dnr #2025-03908-02).

## Data Availability

All data produced will be available online after acceptance in a peer-reviewed journal.

## Acknowledgments

The Norwegian Mother, Father and Child Cohort Study is supported by the Norwegian Ministry of Health and Care Services and the Ministry of Education and Research. We are grateful to all the participating families in Norway who take part in this on-going cohort study. We thank the Norwegian Institute of Public Health (NIPH) for generating high-quality genomic data. This research is part of the HARVEST collaboration, supported by the Research Council of Norway (#229624). We also thank the NORMENT Centre for providing genotype data, funded by the Research Council of Norway (#223273), South-East Norway Health Authorities and Stiftelsen Kristian Gerhard Jebsen. We further thank the Center for Diabetes Research, the University of Bergen for providing genotype data and performing quality control and imputation of the data funded by the ERC AdG project SELECTionPREDISPOSED, Stiftelsen Kristian Gerhard Jebsen, Trond Mohn Foundation, the Research Council of Norway, the Novo Nordisk Foundation, the University of Bergen and the Western Norway Health Authorities. Test was performed using digital laboratories in HUNT Cloud at the Norwegian University of Science and Technology, Trondheim, Norway. We are grateful for outstanding support from the HUNT Cloud community. We are extremely grateful to all the families who took part in this study, the midwives for their help in recruiting them, and the whole ALSPAC team, which includes data collection staff, data and administrations staff, technical managers and the technical staff with the Bristol Bioresource Laboratory, based within the University of Bristol. The UK Medical Research Council and Wellcome (Grant ref: MR/Z505924/1) and the University of Bristol provide core support for ALSPAC. This publication is the work of the authors and P.S.N., B.J. and K.Y. will serve as guarantors for the contents of this paper. A comprehensive list of grants funding is available on the ALSPAC website (http://www.bristol.ac.uk/alspac/external/documents/grant-acknowledgements.pdf); This research was specifically funded by: MRC;MR/M009351/1;Abi Fraser, John Templeton Foundation; Grant ID: 60828; Jean Golding, Wellcome Trust; WT092830/Z/10/Z; Debbie Lawlor, Wellcome Trust;WT088806. Genomewide genotyping data was generated by Sample Logistics and Genotyping Facilities at Wellcome Sanger Institute and LabCorp (Laboratory Corporation of America) using support from 23andMe.

## Author contributions

Conceptualization: K.Y. and P.S.N. Data curation: P.S.N., M.V., E.C.C. and K.Y. Formal analysis: K.Y., P.S.N., H.S., J.J. and X.W. Funding acquisition: P.S.N., B.J., P.R.N. and A.H. Investigation: K.Y. and P.S.N. Methodology: K.Y., P.S.N., J.J., H.S., X.W. and M.V. Project administration: K.Y., P.S.N. and B.J. Resources: B.J., S.J., P.R.N., E.C.C., O.A.A., P.M. and A.H. Software: K.Y., P.S.N., H.S. and X.W. Supervision: P.S.N., B.J. and S.N. Validation: K.Y., P.S.N., J.J., H.S. and X.W. Visualization: K.Y., P.S.N. and X.W. Writing – original draft: K.Y. and P.S.N. Writing – review & editing: K.Y., P.S.N., J.J., H.S., X.W., S.N., B.J., S.J., M.V., P.R.N., E.C.C., O.A.A., P.M., A.H.

## Data and materials availability

Due to sensitive information, access to raw data requires an application to the administrative board of the Norwegian Mother, Father and Child Cohort Study, conditioned on an approved ethical application. Summary statistics from the GWASs created in this project (maternal and fetal GWASs on gestational duration and birth weight in whole study population and stratified by parity) are available from the GWAS Catalogue, accession IDs: GCST90824124, GCST90824125, GCST90824126, GCST90824127, GCST90824128, GCST90824129, GCST90824130, GCST90824131, GCST90824132, GCST90824133, GCST90824134, and GCST90824135. The weights for constructing the polygenic scores of gestational duration in nulliparous women, multiparous women, and the overall study population are available at the PGS Catalog under publication ID PGP000810 and score IDs PGS012573– PGS012575.

The informed consent obtained from ALSPAC (Avon Longitudinal Study of Parents and Children) participants does not allow the data to be made available through any third party maintained public repository. Supporting data are available from ALSPAC on request under the approved proposal number, B5316. Full instructions for applying for data access can be found here: http://www.bristol.ac.uk/alspac/researchers/access/. The ALSPAC study website contains details of all available data (http://www.bristol.ac.uk/alspac/researchers/our-data/).

Analyses were executed with Ubuntu 20.04.4, Long Term Support, and analysis workflow was managed using Snakemake (53), in a defined Conda environment (54). The code can be accessed at Zendo https://doi.org/10.5281/zenodo.18833058 or GitHub repository: https://github.com/PerinatalLab/Parity_gd_bw.

