## Supplementary Material for "Nulliparity modifies the maternal genetic effects on gestational duration but not on birth weight"

Science Advances
AAAS

### Supplementary Materials for

#### Nulliparity modifies the maternal genetic effects on gestational duration but not on birth weight

Karin Ytterberg *et al.*

Corresponding author:
Pol Solé-Navais,;
Bo Jacobsson,;
Karin Ytterberg,

**This PDF file includes:**

Supplementary Text
Figs. S1 to S16
Tables S1 to S18 captions

**Other Supplementary Materials for this manuscript include the following:**
Tables S1 to S18

References (see main text)

**Supplementary Text**

Supplementary Note 1. Fetal gestational duration GWAS whole population

One of the identified loci have previously been reported (*IL1A/B*). The lead variant at the *LSM4* locus (rs4808779, beta = -0.39 days, p-value = $2\times 10^{-9}$, MAF = 33.8%) is fine-mapped as an expression-QTL for *JUND* and *IQCN* genes in brain and immune cell types, respectively, with concordant effect direction (i.e., the allele associated with higher gestational duration is associated with higher gene expression). *JUND* is a transcription factor that has been suggested to play a role in the switch from quiescent to contractile phenotype in the human pregnant myometrium (55). This contrasts with the fact that the discovery has been performed using the fetal genome, highlighting the difficulties in teasing out causal genes in phenotypes shaped by maternal and fetal genomes.

Supplementary Note 2. Random forest models - adding known environmental factors

Across all phenotypes and all genomes, the prediction accuracy was higher when including both genetic and environmental features compared to including only genetic variants, except for gestational duration on the fetal genome in pregnancies of nulliparous woman where there was no difference between the two feature sets (Table S15). The trait with the lowest improvement in prediction accuracy when including both genetic and environmental features was gestational duration in the maternal genome (Table S15). This may indicate that gestational duration has an unresolved complexity compared to birth weight. In the parity-stratified study population, the predictive accuracy of the maternal genome on gestational duration was also higher in nulliparous woman, following the same pattern as when using only genetic features (Table S15).

Supplementary Methods

To optimize the performance of the random forest the following hyperparameters were tuned: number of decision trees (n trees), the ratio of features that were available to be considered at each split (mtry ratio), minimum number of observations in a leaf node (min node size) and mechanism used for node splitting (splitrule). In addition, subsampling without replacement (replace) of different fractions (sample fraction) was also tested instead of replacement (bootstrap sampling), since the out-of-bag error in random forests can overestimate the actual test error (56, 57). The search space used for hyperparameters were:

- n trees: Any integer between 20 and 1000
- mtry ratio: Any float between 0 and 1
- min node size: Any integer between 1 and 40
- splitrule: "variance", "extratrees" and "maxstat"
- replace: True or False
- Sample fraction: Any float between 0.1 and 1

Tuning was done using Bayesian optimization. The set of hyperparameters with the highest r-squared out-of-bag-score was selected for each model (Table M1).

The same set of data is used both for tuning hyperparameters and evaluating model performance. The out-of-bag samples are selected randomly which means that it is possible that the samples used for testing and selecting hyperparameters are used again for testing when evaluating the model performance, thus causing an overestimation of the model performance due to overfitted hyperparameters. While running the evaluation multiple times reduces the risk of this having any effect on the overall results, we wanted to be certain that this is the case. We created a new setup using nested cross-validation. This method is more time consuming and computationally heavier, but it ensures that the test set used for evaluating the model has never been seen by the model. We used this method on a subset of the different models (gestational duration maternal genome for both feature sets) and compared the results to those obtained with our original setup. As seen in the Table M2 the difference in model performance is very small, suggesting that the risk of overestimating model performance is very small.

**Table M1** – Optimal hyperparameters for the random forest models

| Model description | | | | Hyperparameters | | | | | |
|---|---|---|---|---|---|---|---|---|---|
| **Features[a]** | **Phenotype[b]** | **Genome** | **Parity** | **n trees** | **mtry ratio** | **min node size** | **splitrule** | **replace** | **Sample fraction** |
| G | gd | Maternal | Whole population | 669 | 0.253 | 40 | extratrees | TRUE | 0.444 |
| G | bw | Maternal | Whole population | 854 | 0.541 | 40 | extratrees | TRUE | 0.176 |
| G | gd | Fetal | Whole population | 409 | 0.377 | 38 | maxstat | TRUE | 0.455 |
| G | bw | Fetal | Whole population | 834 | 0.783 | 1 | variance | FALSE | 0.113 |
| G | gd | Maternal | Nulliparous | 669 | 0.253 | 40 | extratrees | TRUE | 0.444 |
| G | bw | Maternal | Nulliparous | 889 | 0.730 | 22 | extratrees | FALSE | 0.308 |
| G | gd | Fetal | Nulliparous | 61 | 0.583 | 2 | maxstat | TRUE | 0.204 |
| G | bw | Fetal | Nulliparous | 725 | 0.974 | 39 | extratrees | FALSE | 0.305 |
| G | gd | Maternal | Multiparous | 825 | 0.902 | 11 | maxstat | TRUE | 0.479 |
| G | bw | Maternal | Multiparous | 940 | 0.811 | 17 | extratrees | TRUE | 0.222 |
| G | gd | Fetal | Multiparous | 577 | 0.597 | 37 | maxstat | TRUE | 0.978 |
| G | bw | Fetal | Multiparous | 789 | 0.656 | 28 | variance | FALSE | 0.390 |
| GE | gd | Maternal | Whole population | 669 | 0.253 | 40 | extratrees | TRUE | 0.444 |
| GE | bw | Maternal | Whole population | 930 | 0.511 | 10 | extratrees | TRUE | 0.219 |
| GE | gd | Fetal | Whole population | 881 | 0.907 | 40 | maxstat | TRUE | 0.360 |
| GE | bw | Fetal | Whole population | 939 | 0.635 | 24 | extratrees | TRUE | 0.912 |
| GE | gd | Maternal | Nulliparous | 815 | 0.210 | 40 | extratrees | TRUE | 0.369 |
| GE | bw | Maternal | Nulliparous | 629 | 0.886 | 25 | extratrees | TRUE | 0.106 |
| GE | gd | Fetal | Nulliparous | 29 | 0.721 | 34 | maxstat | FALSE | 0.950 |
| GE | bw | Fetal | Nulliparous | 773 | 0.704 | 38 | extratrees | TRUE | 0.992 |
| GE | gd | Maternal | Multiparous | 669 | 0.253 | 40 | extratrees | TRUE | 0.444 |
| GE | bw | Maternal | Multiparous | 580 | 0.807 | 27 | extratrees | TRUE | 0.114 |
| GE | gd | Fetal | Multiparous | 478 | 0.911 | 24 | maxstat | TRUE | 0.318 |
| GE | bw | Fetal | Multiparous | 454 | 0.847 | 27 | extratrees | FALSE | 0.303 |
| G – MT allele only | gd | Maternal | mother-child duos | 556 | 0.963 | 26 | maxstat | TRUE | 0.475 |
| G – MnT allele only | gd | Maternal | mother-child duos | 273 | 0.444 | 14 | maxstat | TRUE | 0.823 |
| G - PT allele only | gd | Maternal | mother-child duos | 678 | 0.0370 | 13 | maxstat | TRUE | 0.859 |

[a] G = genetic features only, GE = genetic and environmental features

[b] gd = gestational duration, bw = birth weight

**Table M2** – Model performance ($r^2$) using Bayesian optimization and nested cross-validation respectively

| **Model description** | | | | **$r^2$ (95% credible interval)** | |
|---|---|---|---|---|---|
| **Features[a]** | **Phenotype[b]** | **Genome** | **parity** | **Bayesian optimization** | **Nested cross-validation** |
| G | gd | Maternal | Whole population | 0.00602 (0.00571 - 0.00632) | 0.00916 (0.00881 - 0.00975) |
| G | gd | Maternal | 0 | 0.00848 (0.00802 - 0.00895) | 0.0121 (0.0118 - 0.0137) |
| G | gd | Maternal | > 0 | 0.00305 (0.00294 - 0.00315) | 0.00671 (0.00577- 0.00754) |
| GE | gd | Maternal | Whole population | 0.00943 (0.00908 – 0.00978) | 0.0120 ( 0.0113 – 0.0123) |
| GE | gd | Maternal | 0 | 0.0108 (0.0103 – 0.0113) | 0.0139 (0.0126 – 0.0148) |
| GE | gd | Maternal | > 0 | 0.00777 (0.00732 – 0.00824) | 0.00951 (0.00872 – 0.00997) |

[a] G = genetic features only, GE = genetic and environmental features

[b] gd = gestational duration, bw = birth weight

**A**

Gestational duration of mothers included in the study

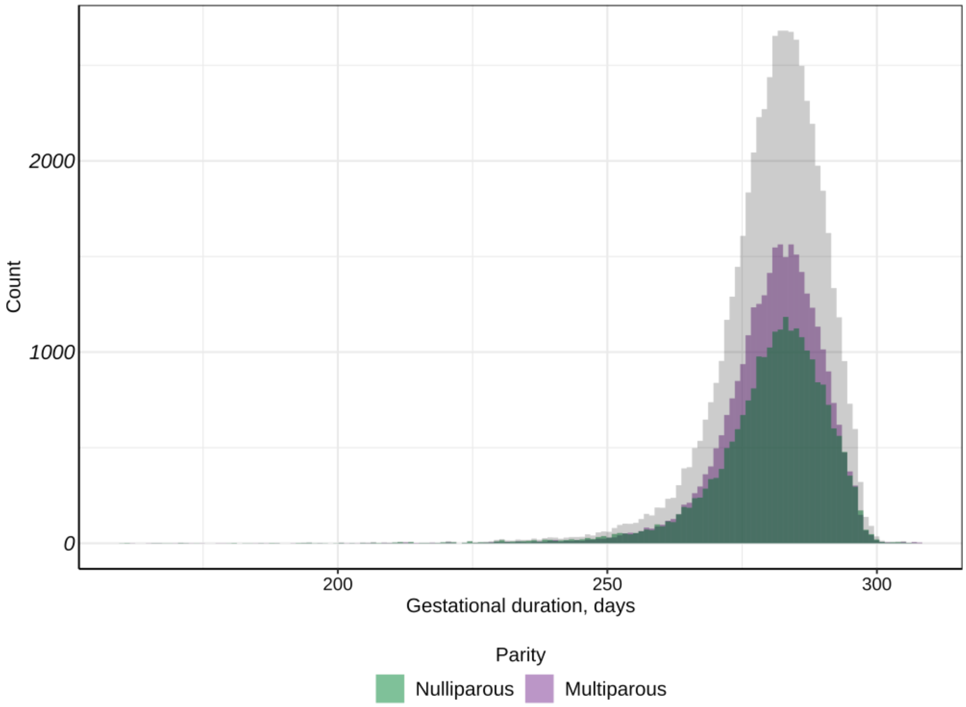


**B**

Gestational duration of children included in the study

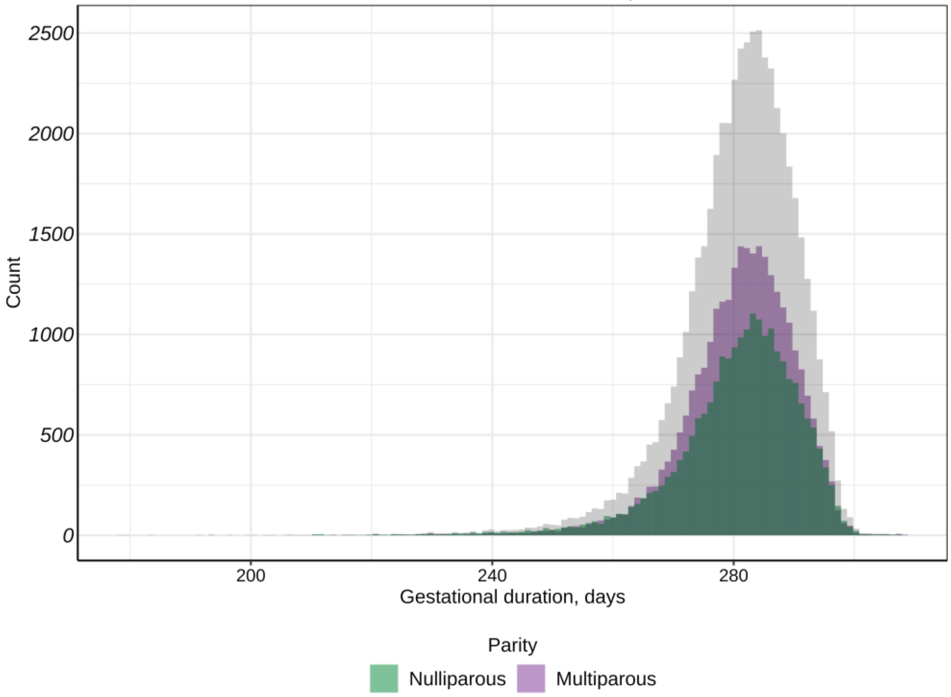

**C**

Offspring birth weight for mothers included in the study

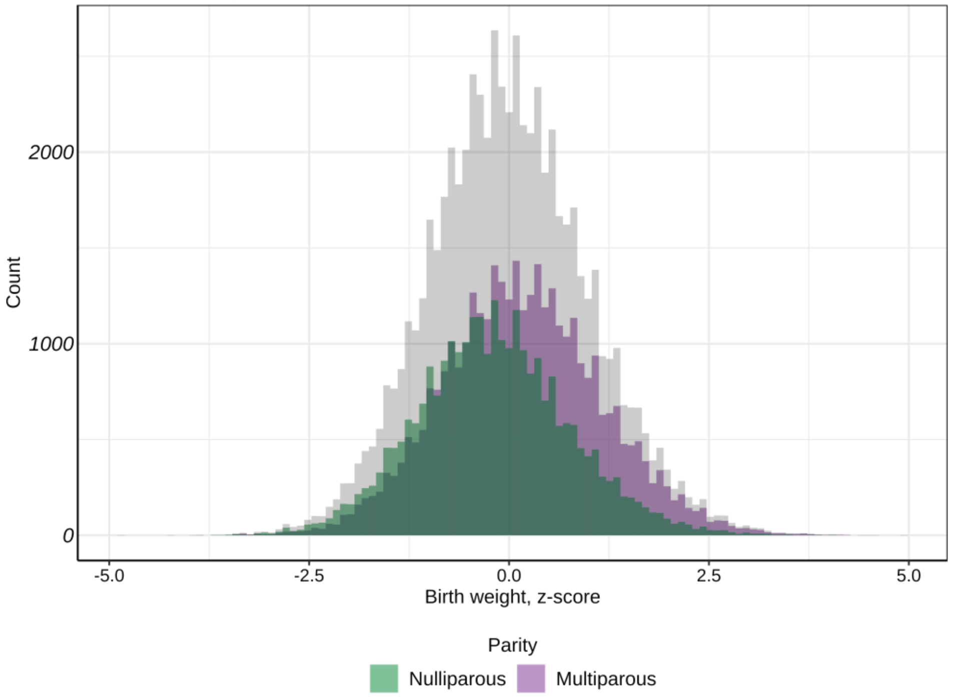


**D**

Birth weight for children included in the study

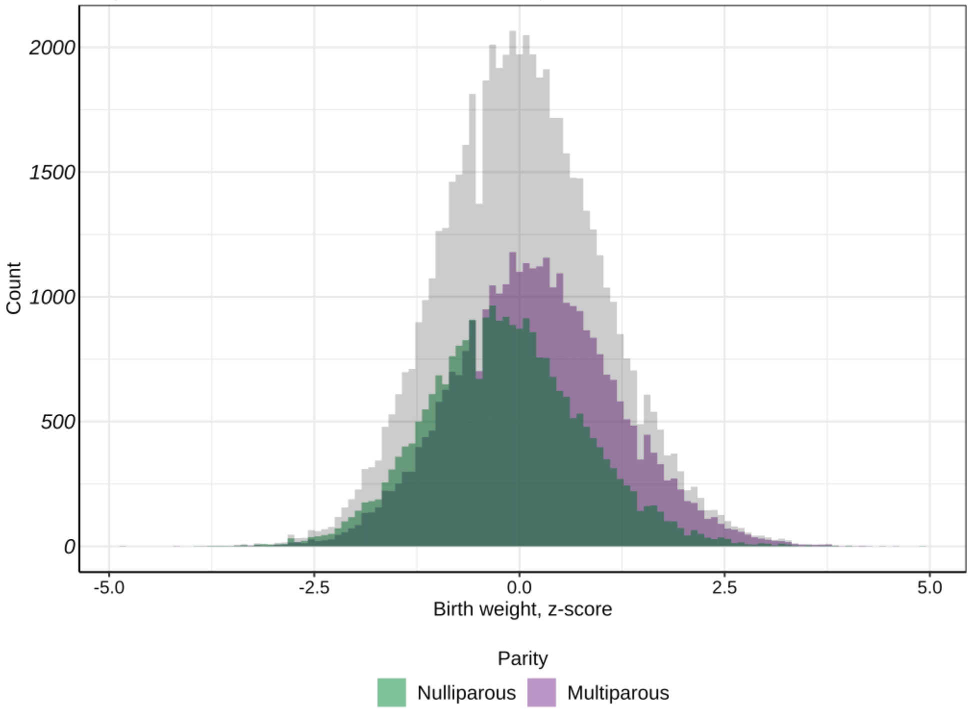


**Fig. S1.** Histogram of the distributions of (A) gestational duration in days for mothers included in the study, (B) gestational duration in days for children included in the study, (C) offspring's birth weight as z-scores for mothers included in the study, and (D) birth weight as z-scores for children included in the study. The histograms are stratified by nulliparous women (green) and multiparous women (purple), and the grey distribution represents the whole population.

**A**

Nulliparous women, maternal genome, gestational duration

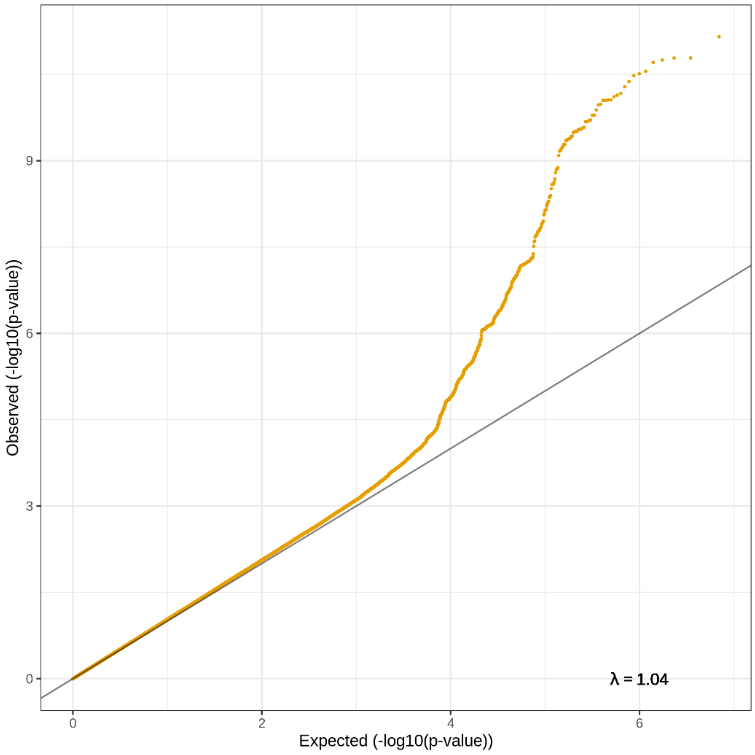


**B**

Multiparous women, maternal genome, gestational duration

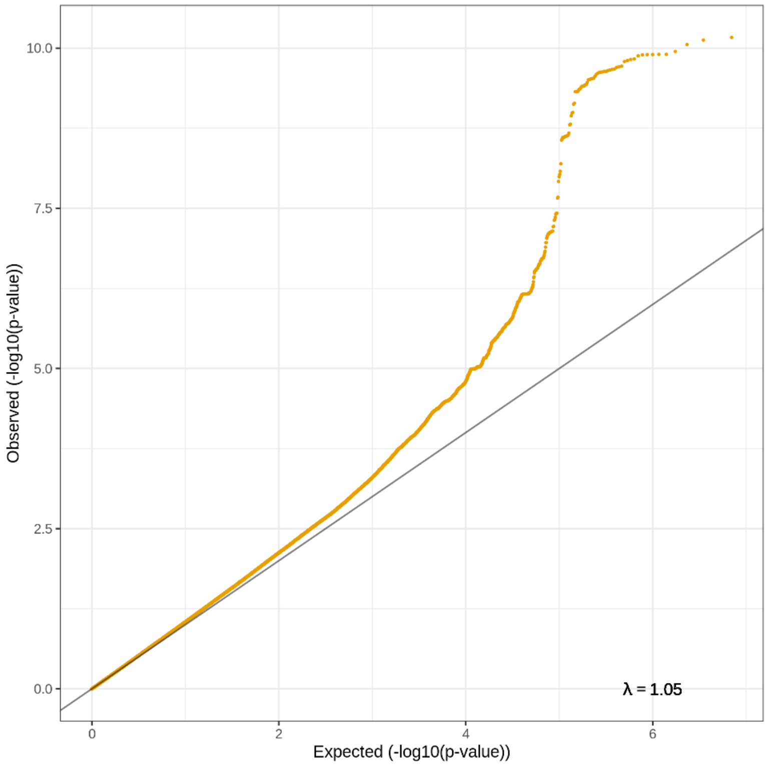

**C**

Whole study population maternal genome gestational duration

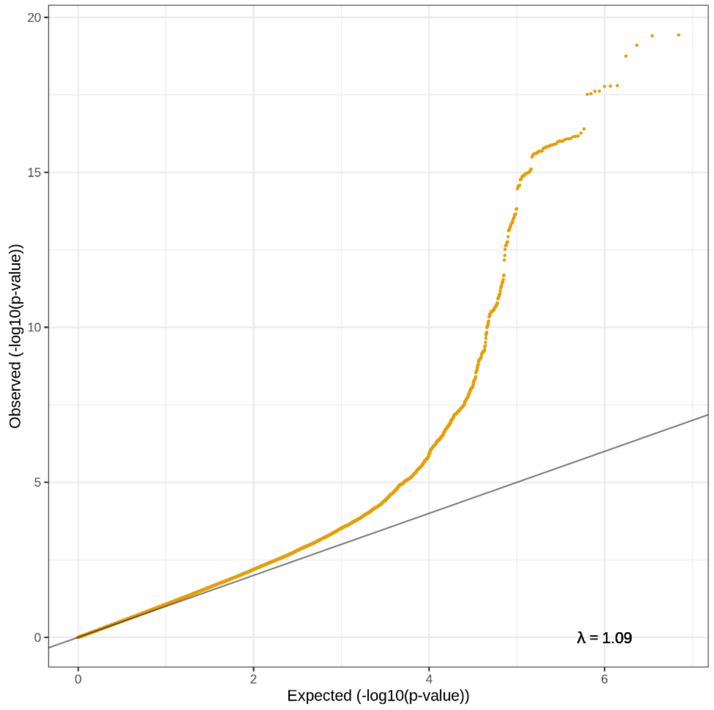


**D**

Whole study population maternal genome gestational duration

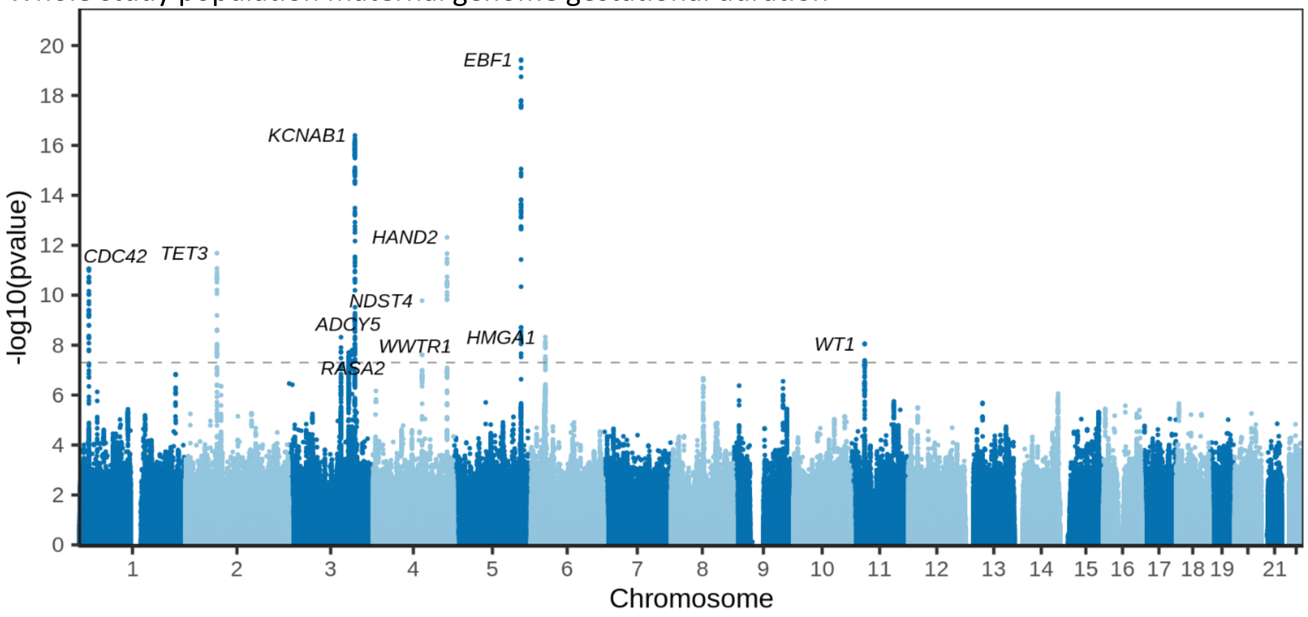


**Fig. S2.** QQ-plots and a Manhattan plot of the gestational duration genome-wide association studies on the maternal genome. The QQ-plots shown for (A) nulliparous women, (B) multiparous women and (C) the whole study population include the lambda genomic control and demonstrate little to no test statistic inflation. In (D), the Manhattan plot for the whole study population, the dashed line represents the genome-wide significant level (p-value = 5 x $10^{-8}$). The x-axis shows the chromosome position and y-axis the two-sided p-value of the GWAS. Variants reaching the significance threshold have the nearest protein-coding gene displayed.

A

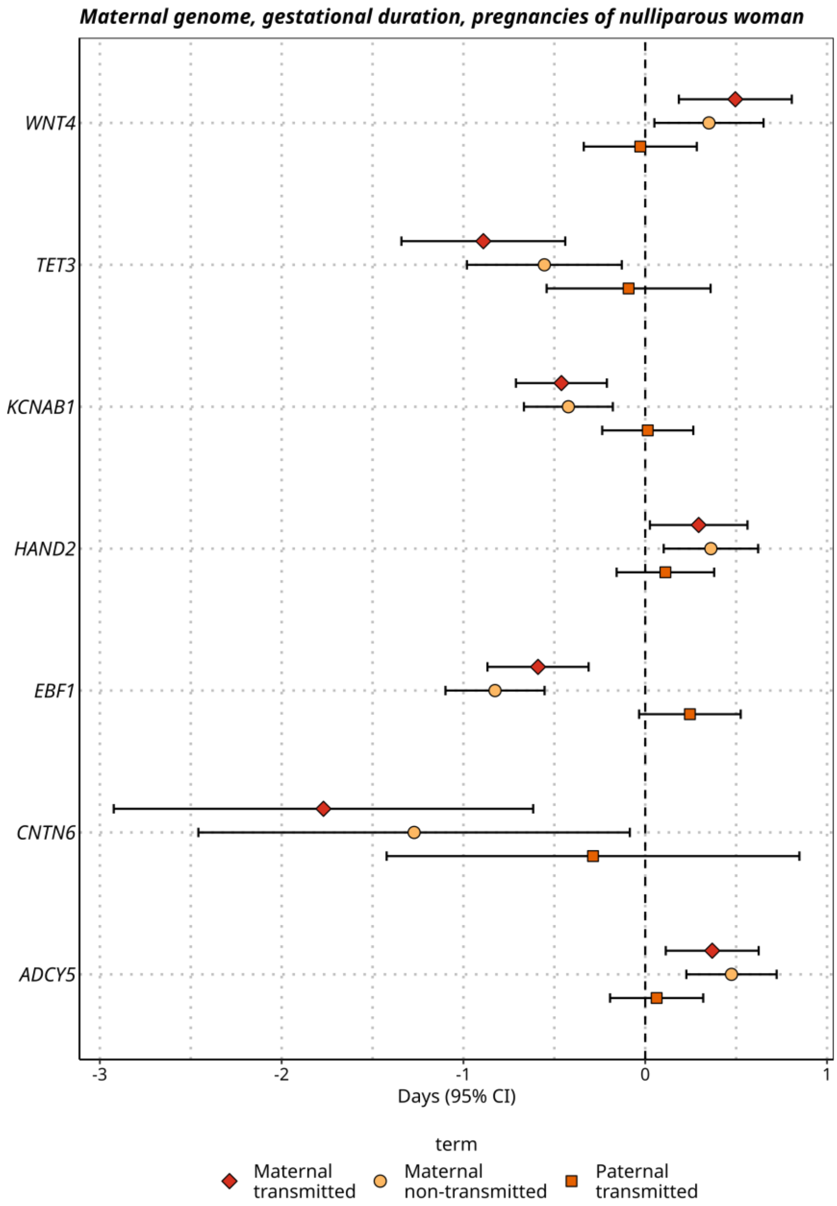
Maternal genome, gestational duration, pregnancies of nulliparous woman
WNT4
TET3
KCNAB1
HAND2
EBF1
CNTN6
ADCY5
-3
-2
-1
0
1
Days (95% CI)
term
Maternal transmitted
Maternal non-transmitted
Paternal transmitted

**B**

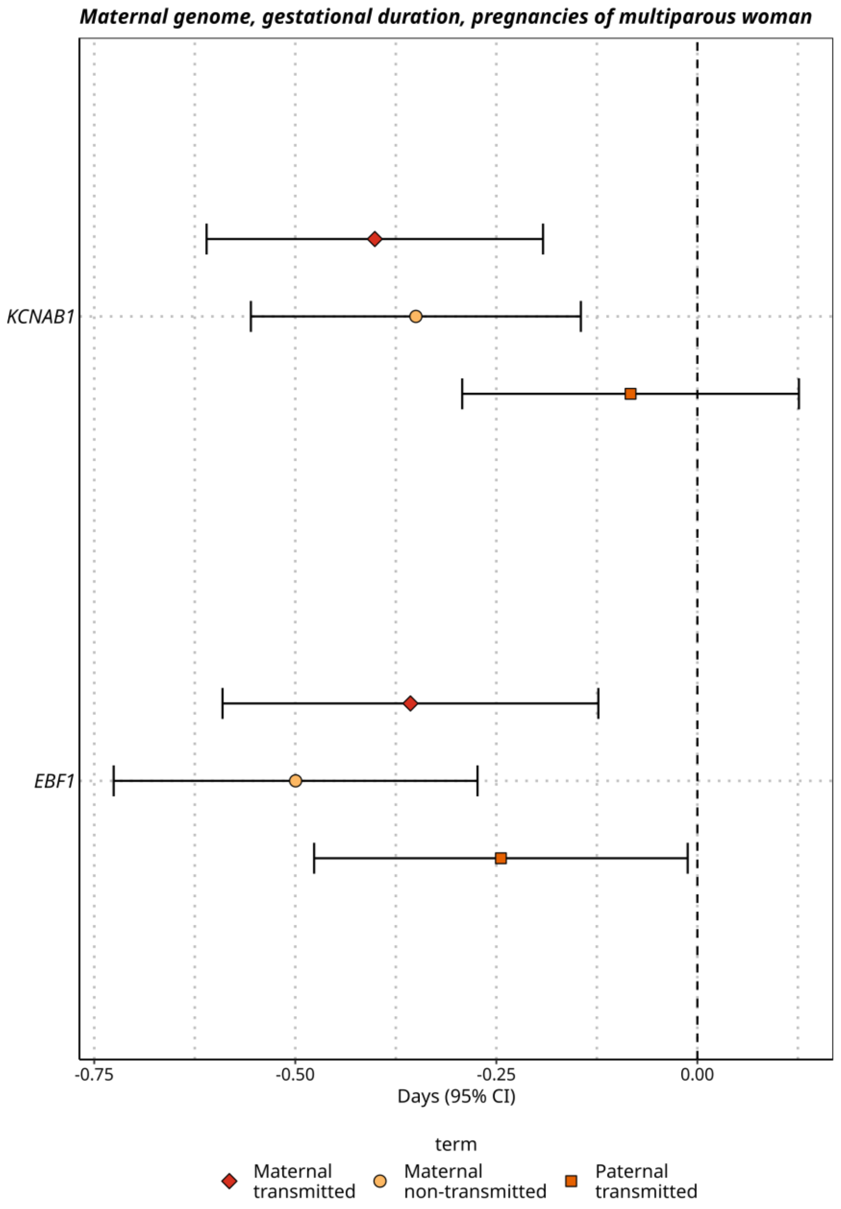
Maternal genome, gestational duration, pregnancies of multiparous woman
KCNAB1
EBF1
-0.75
-0.50
-0.25
0.00
Days (95% CI)
term
Maternal transmitted
Maternal non-transmitted
Paternal transmitted

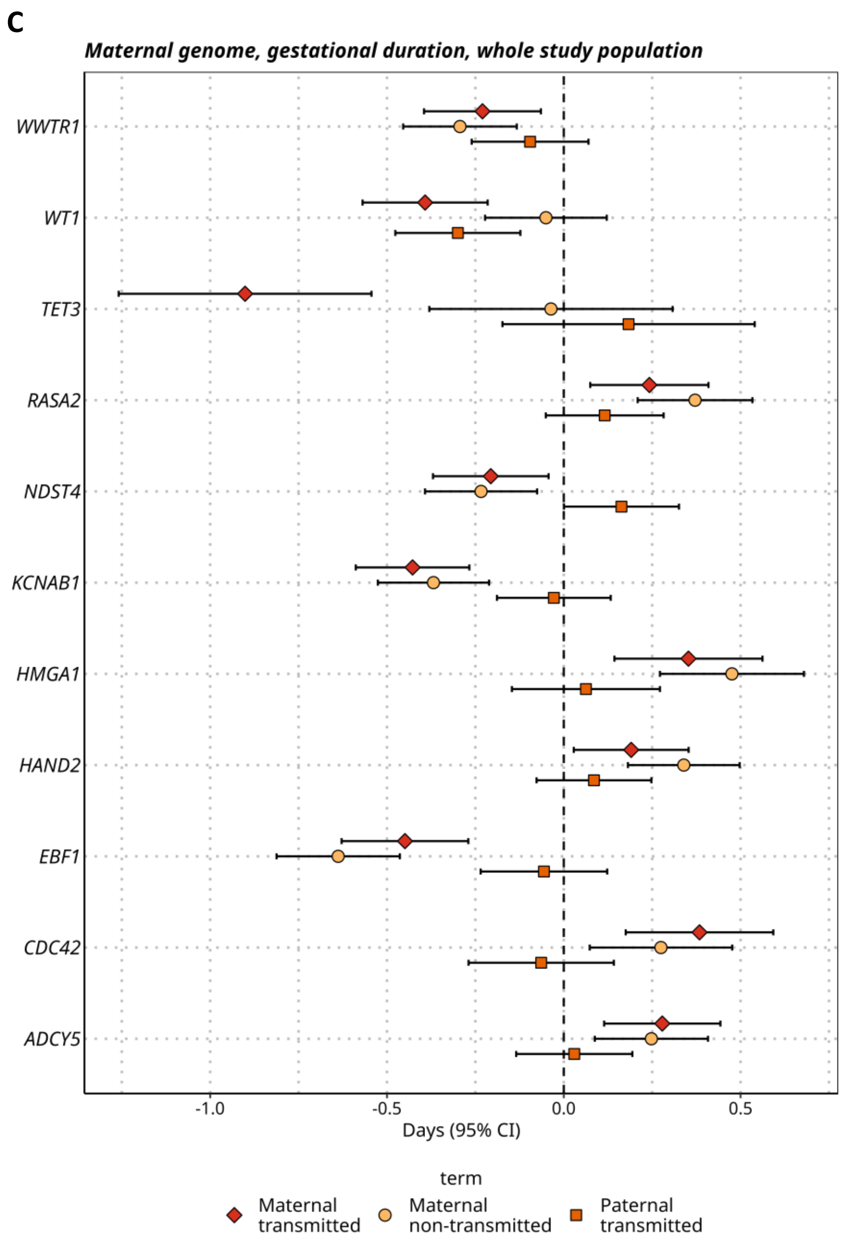


**Fig. S3.** The effect origin using mother-child duos for SNPs reaching the genome wide significance threshold in (A) gestational duration maternal genome pregnancies of nulliparous woman ($n_{duos}$=17,869), (B) gestational duration maternal genome pregnancies of multiparous woman ($n_{duos}$=23,426), (C) gestational duration maternal genome whole study population ($n_{duos}$=41,295). Variants are displayed as the nearest protein-coding gene on the y-axis.

**A**

Pregnancies of nulliparous women, fetal genome, gestational duration

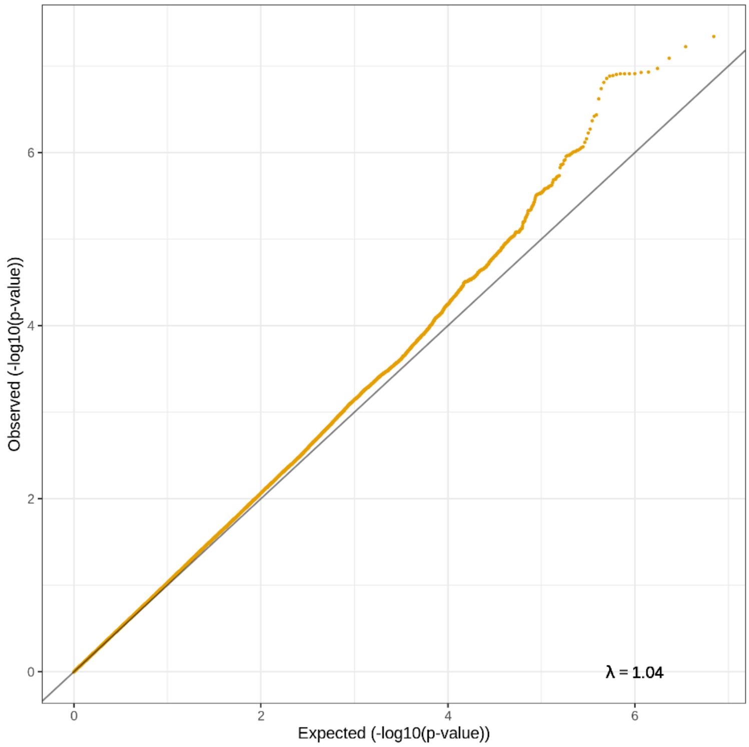


**B**

Pregnancies of multiparous women, fetal genome, gestational duration

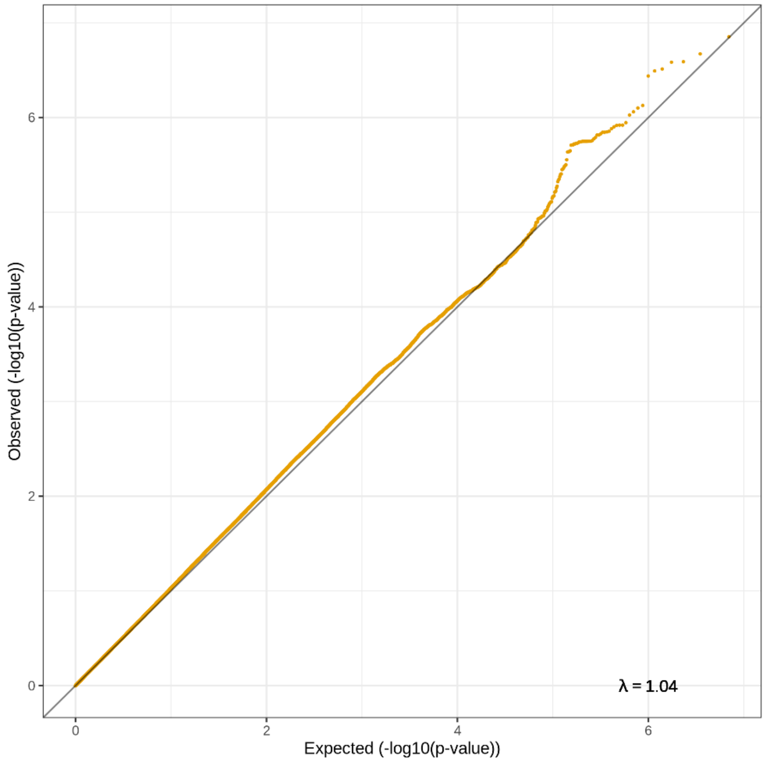

**C**

Whole population fetal genome gestational duration

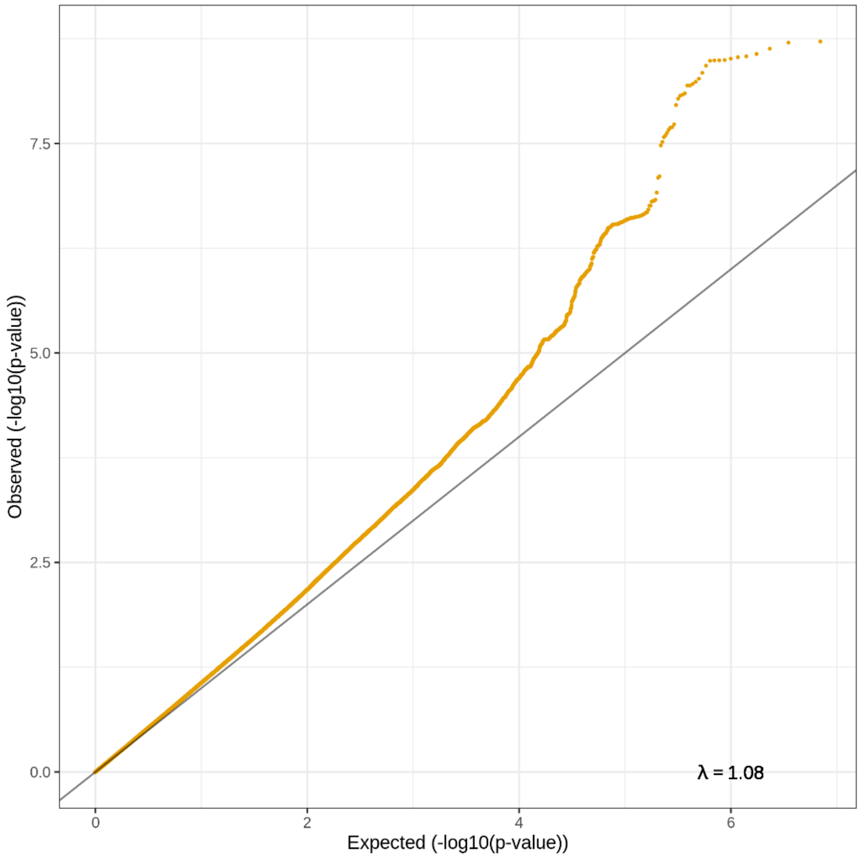


**D**

Pregnancies of nulliparous women (top) and multiparous women (bottom), fetal genome, gestational duration

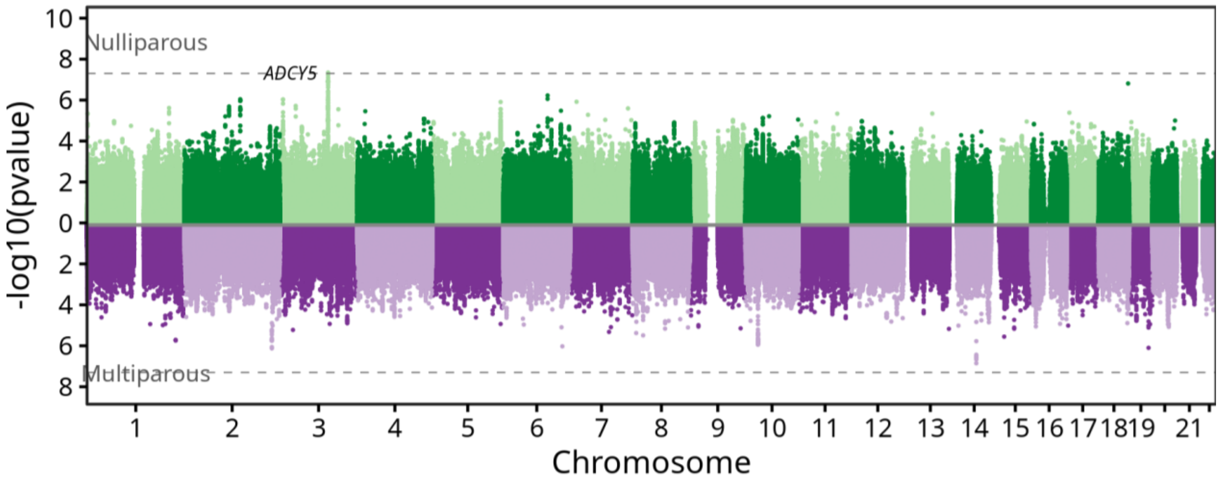

**E**

Whole study population fetal genome gestational duration

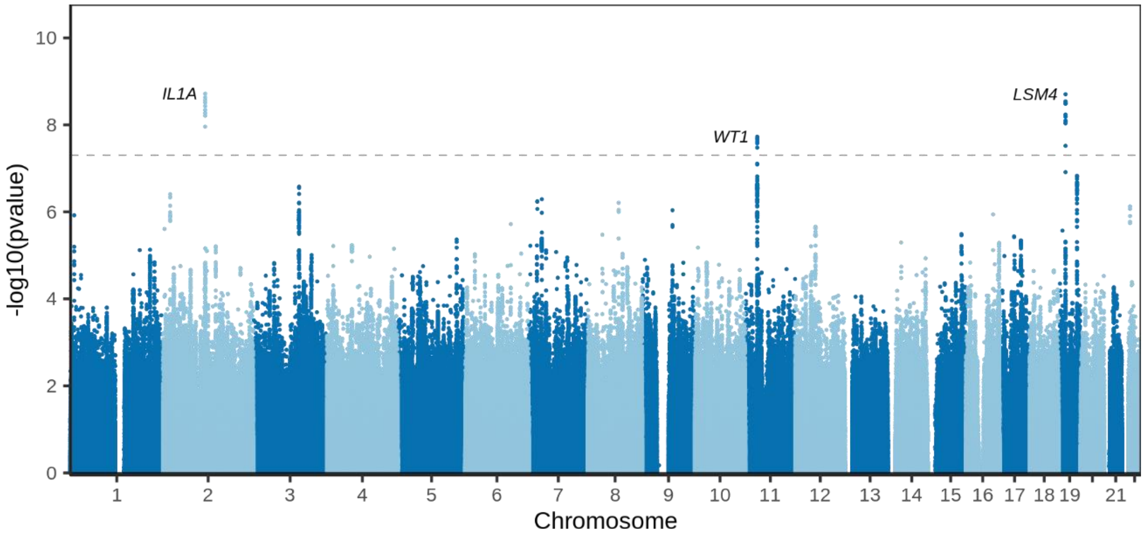


**Fig. S4.** QQ-plots and Manhattan plots of gestational duration genome-wide association studies on the fetal genome. The QQ-plots shown for (A) nulliparous women, (B) multiparous women, and (C) the whole study population include the lambda genomic control and demonstrate little to no test statistic inflation. In the Miami plot for (D) Pregnancies of nulliparous women (top, green) and multiparous women, (bottom, purple), and the Manhattan plot for (E) the whole study population, the dashed line represents the genome-wide significant level (p-value = 5 x $10^{-8}$). The x-axis shows the chromosome position and y-axis the two-sided p-value of the GWAS. Variants reaching the significance threshold have the nearest protein-coding gene displayed.

A

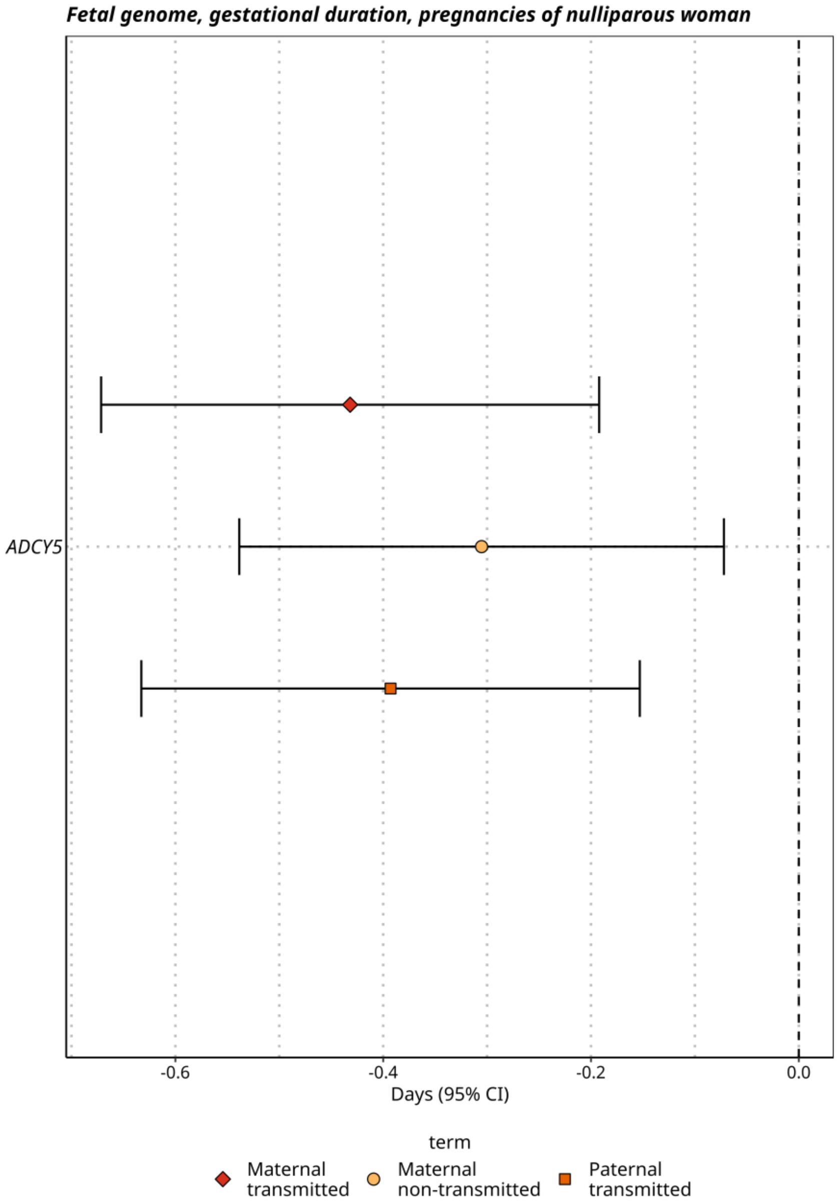

Fetal genome, gestational duration, pregnancies of nulliparous woman
ADCY5
-0.6
-0.4
-0.2
0.0
Days (95% CI)
term
Maternal transmitted
Maternal non-transmitted
Paternal transmitted

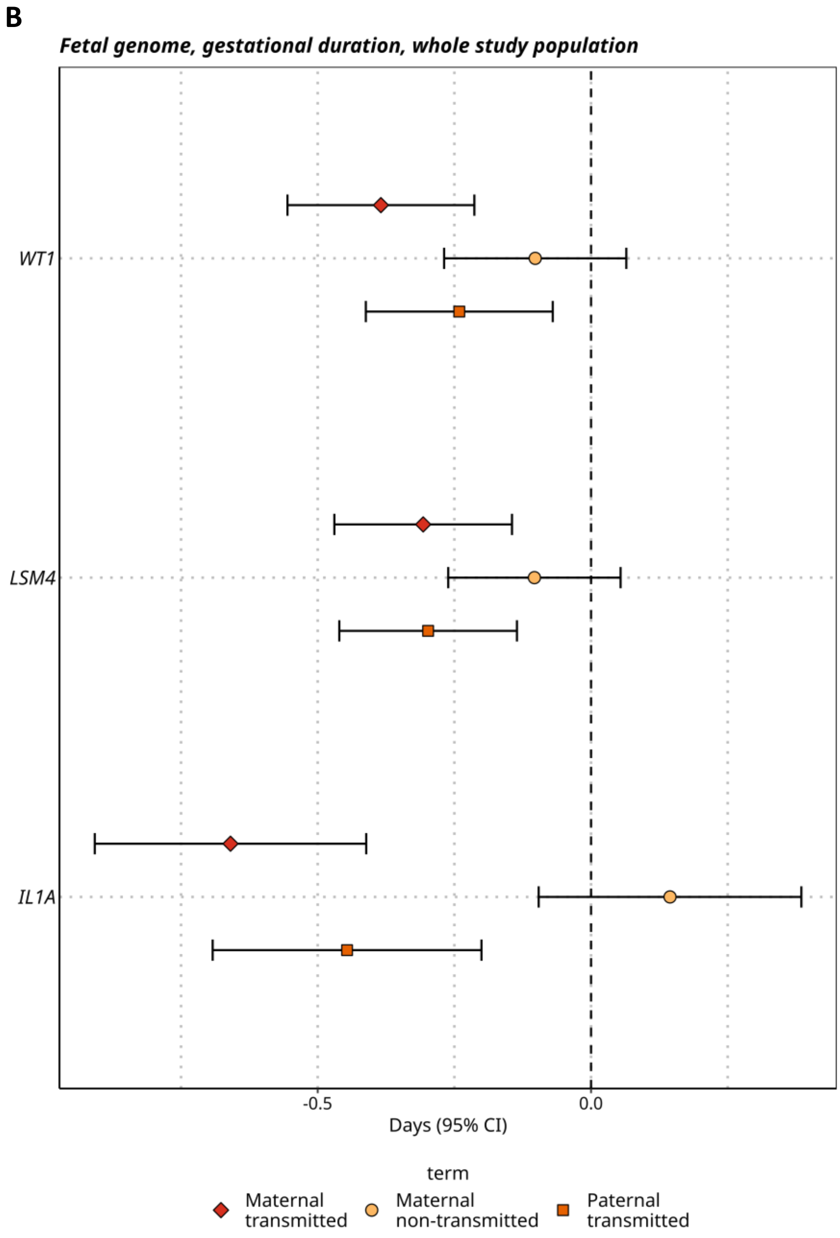


**Fig. S5.** The effect origin using mother-child duos for SNPs reaching the genome wide significance threshold in (A) gestational duration fetal genome pregnancies of nulliparous woman ($n_{duos}$=17,878), (B) gestational duration fetal genome whole study population ($n_{duos}$=40,833). Variants are displayed as the nearest protein-coding gene on the y-axis.

**A**

Nulliparous women, maternal genome, birth weight (z-score)

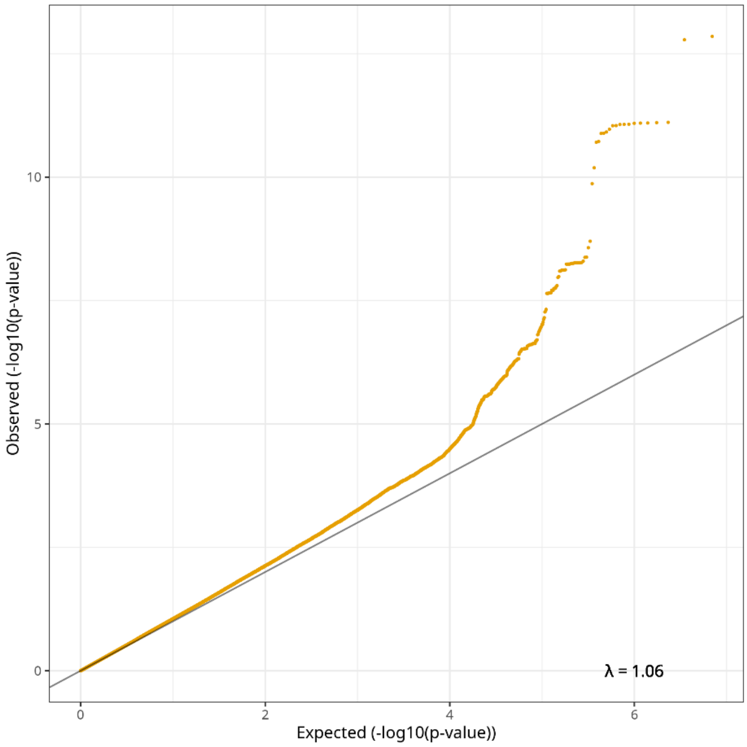


**B**

Multiparous women, maternal genome, birth weight (z-score)

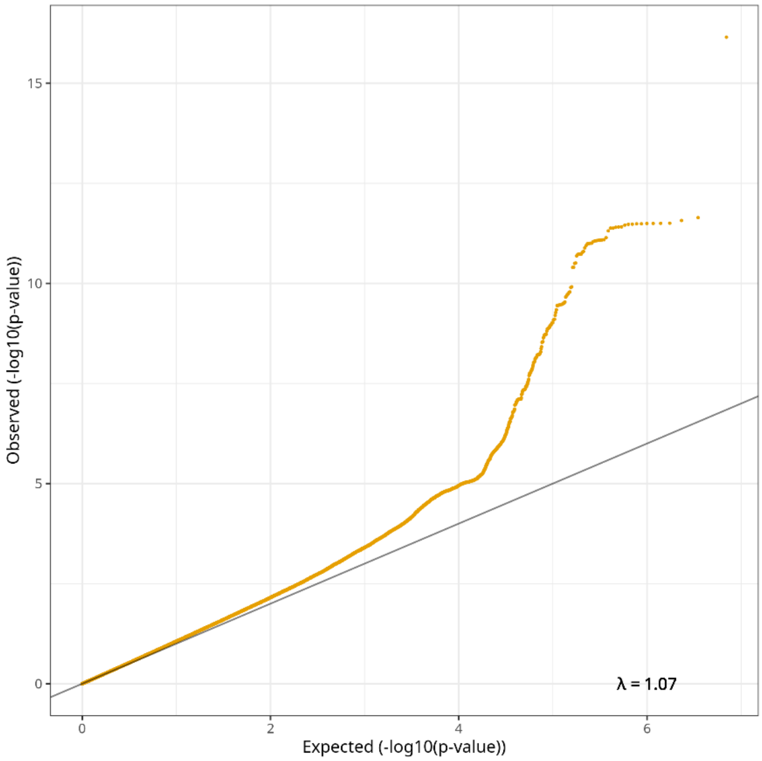

**C**

Whole population maternal genome birth weight (z-score)

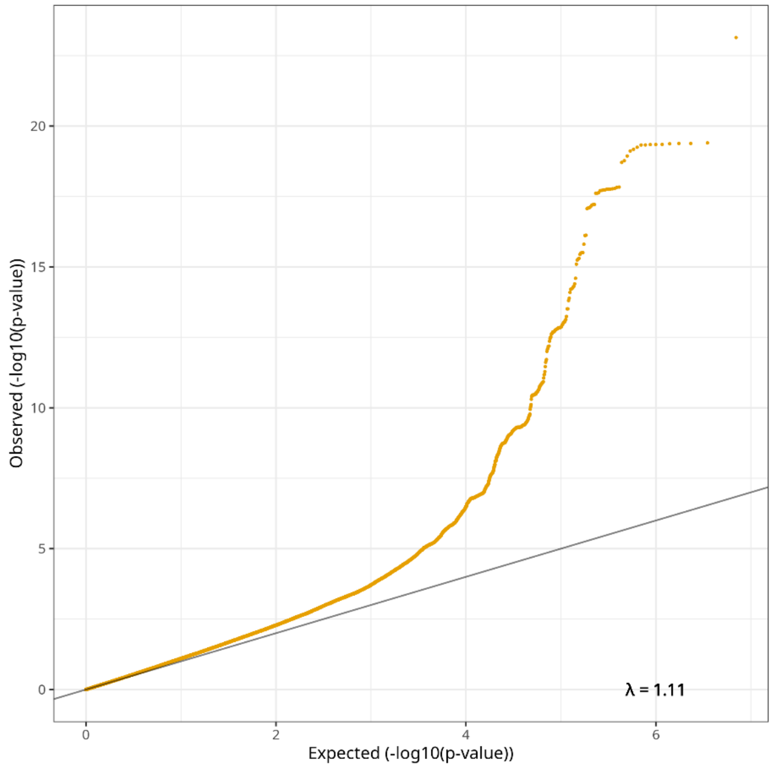


**D**

Pregnancies of nulliparous women (top) and multiparous women (bottom), maternal genome birth weight (z-score)

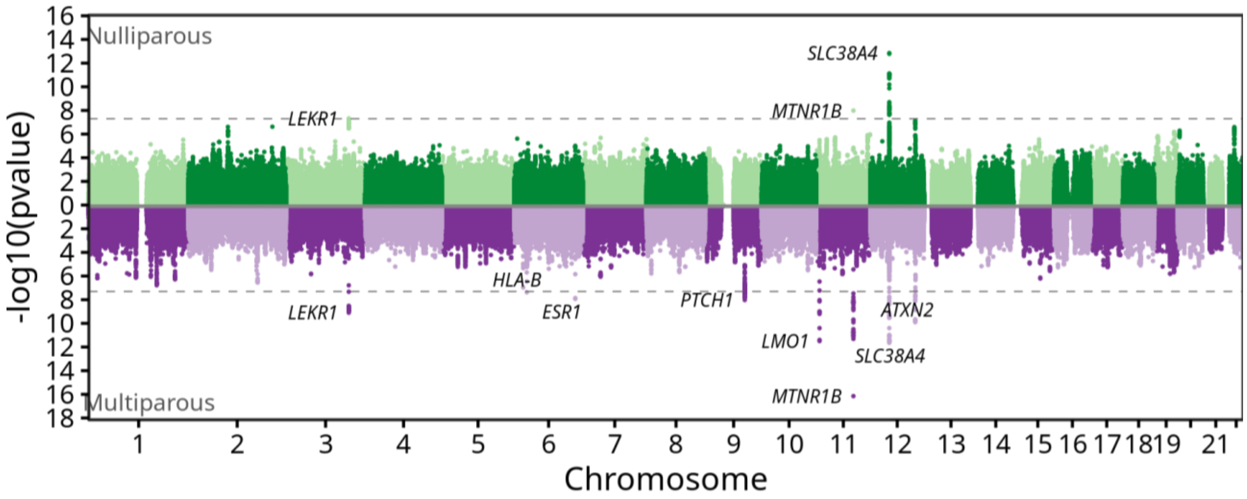

**E**

Whole study population maternal genome birth weight (z-score)

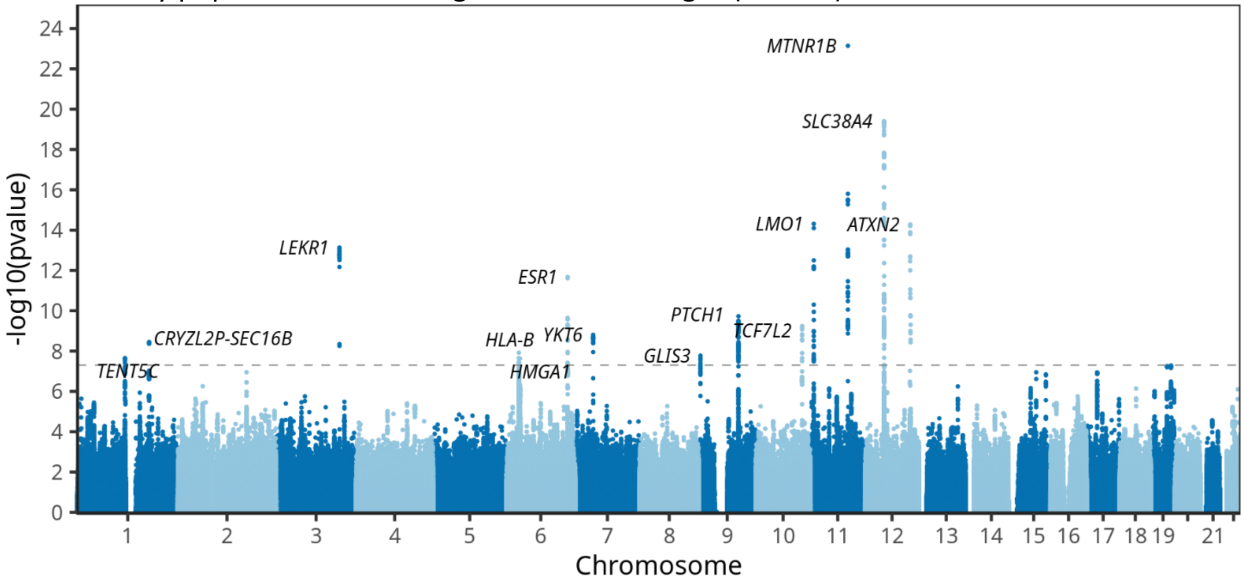


**Fig. S6.** QQ-plots and Manhattan plots of birth weight (z-score) genome-wide association studies on the maternal genome. The QQ-plots shown for (A) nulliparous women, (B) multiparous women, and (C) the whole study population include the lambda genomic control and demonstrate little to no test statistic inflation. In the Miami plot for (D) Pregnancies of nulliparous women (top, green) and multiparous women, (bottom, purple), and the Manhattan plot for (E) the whole study population, the dashed line represents the genome-wide significant level (p-value = 5 x $10^{-8}$). The x-axis shows the chromosome position and y-axis the two-sided p-value of the GWAS. Variants reaching the significance threshold have the nearest protein-coding gene displayed.

**A**

Pregnancies of nulliparous women, fetal genome, birth weight (z-score)

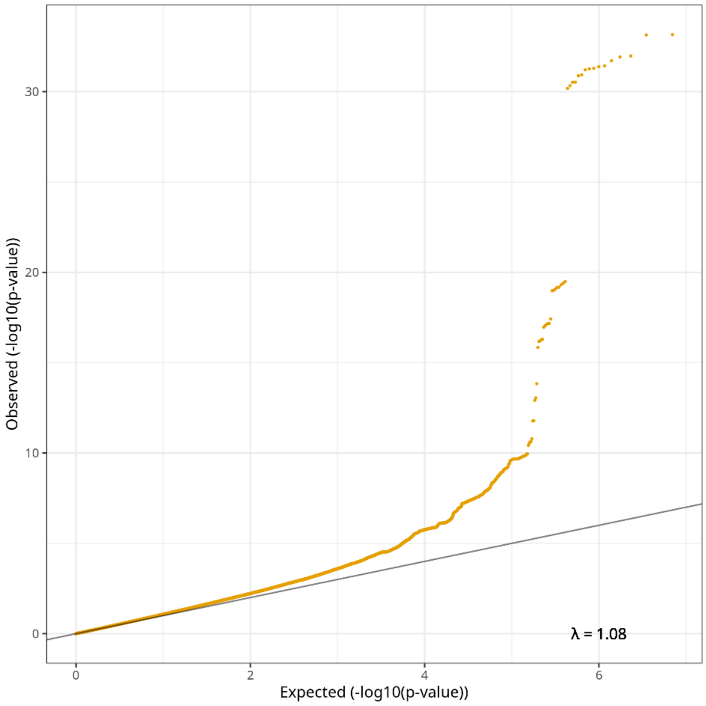


**B**

Pregnancies of multiparous women, fetal genome, birth weight (z-score)

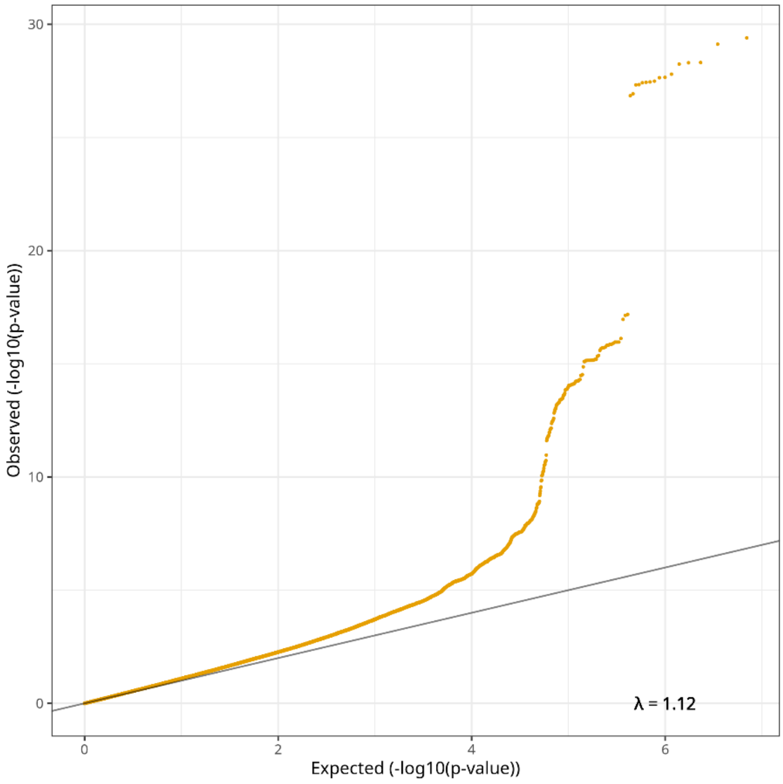

**C**

Whole population, fetal genome, birth weight (z-score)

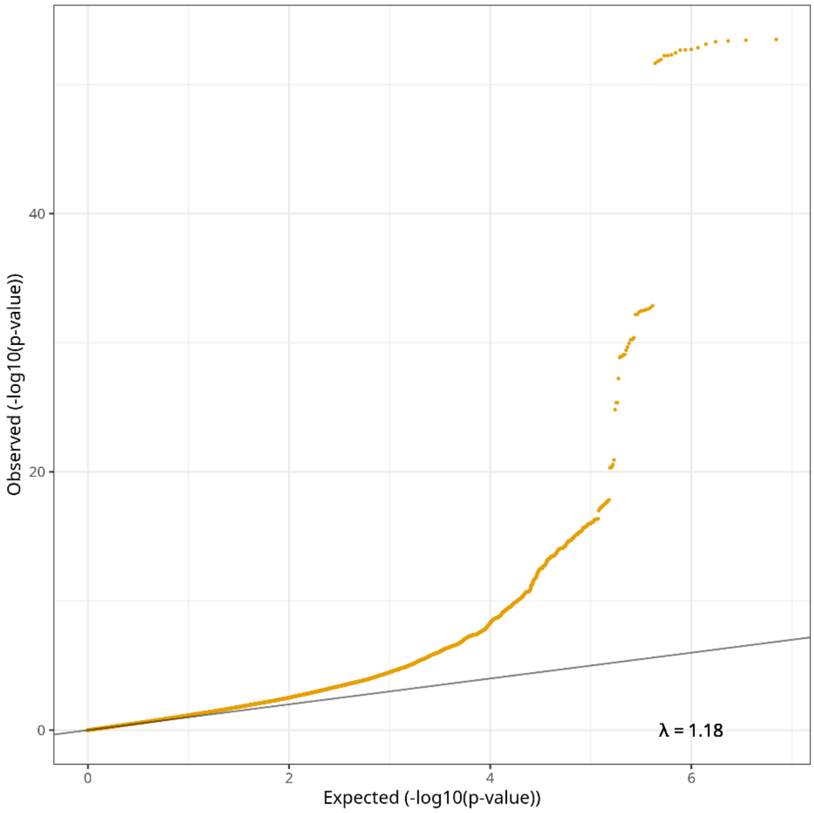


**D**

Pregnancies of nulliparous women (top) and multiparous women (bottom), fetal genome birth weight (z-score)

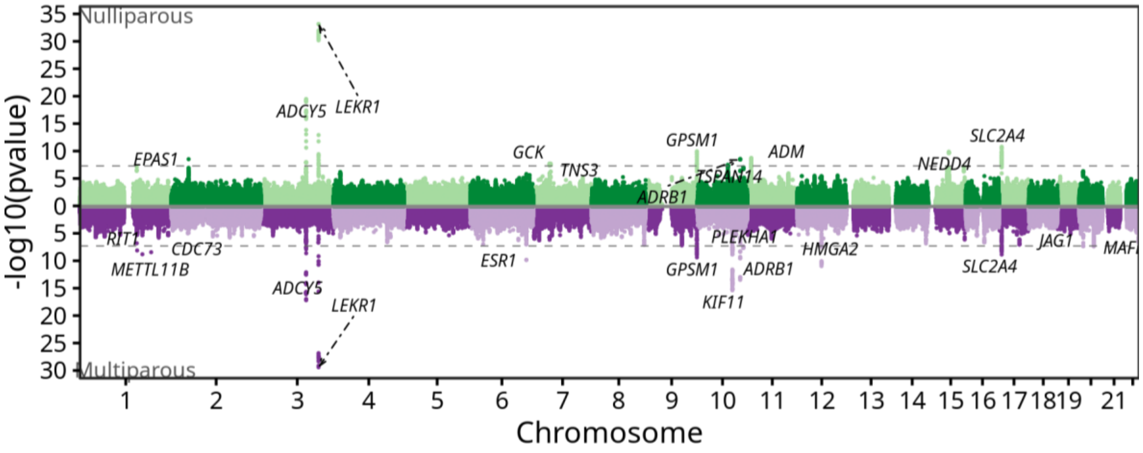

**E**

Whole study population fetal genome birth weight (z-score)

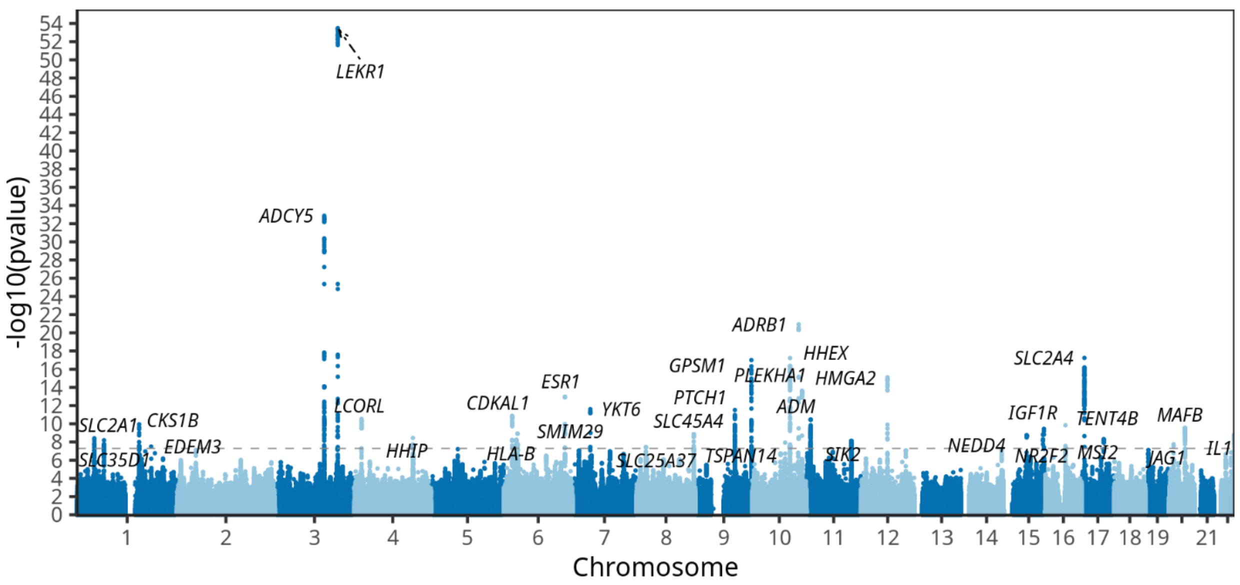


**Fig. S7.** QQ-plots and Manhattan plots of birth weight (z-score) genome-wide association studies on the fetal genome. The QQ-plots shown for (A) nulliparous women, (B) multiparous women, and (C) the whole study population include the lambda genomic control and demonstrate little to no test statistic inflation. In the Miami plot for (D) Pregnancies of nulliparous women (top, green) and multiparous women, (bottom, purple), and the Manhattan plot for (E) the whole study population, the dashed line represents the genome-wide significant level (p-value = 5 x $10^{-8}$). The x-axis shows the chromosome position and y-axis the two-sided p-value of the GWAS. Variants reaching the significance threshold have the nearest protein-coding gene displayed.

**A**

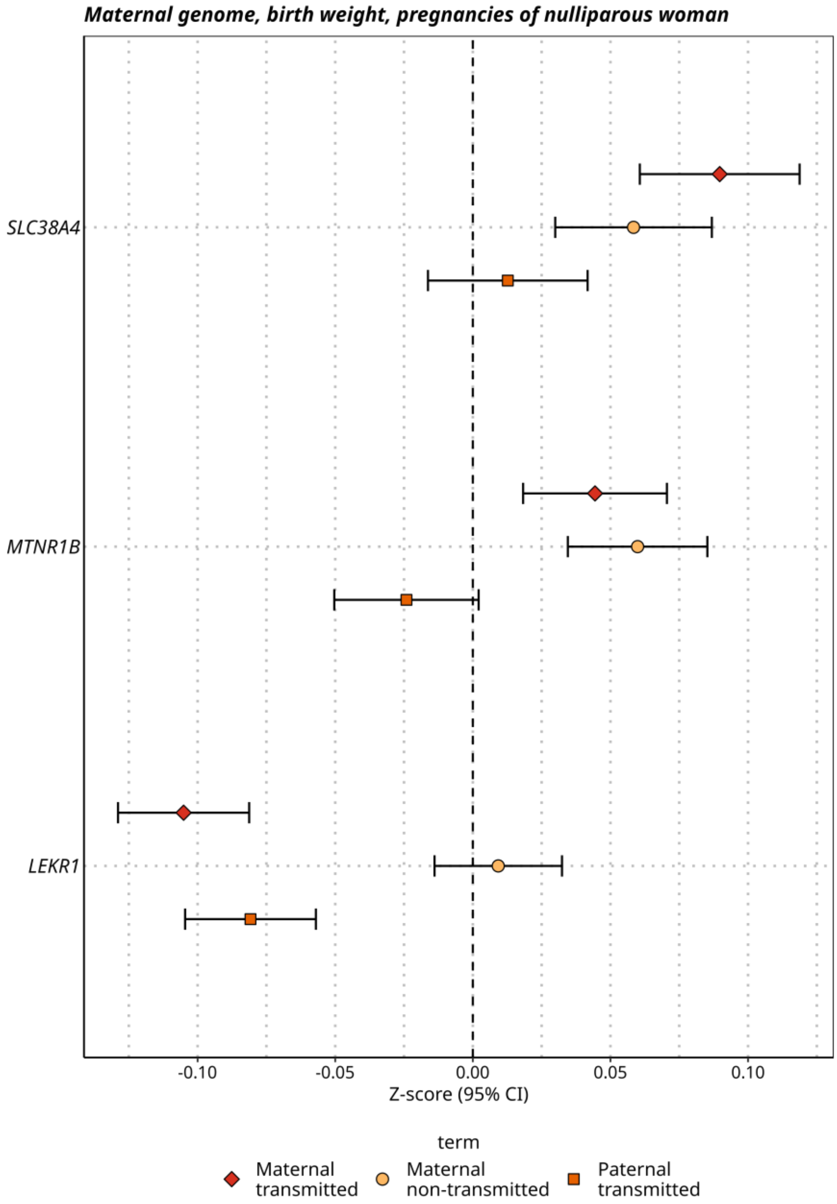
Maternal genome, birth weight, pregnancies of nulliparous woman
SLC38A4
MTNR1B
LEKR1
-0.10
-0.05
0.00
0.05
0.10
Z-score (95% CI)
term
Maternal transmitted
Maternal non-transmitted
Paternal transmitted

**B**

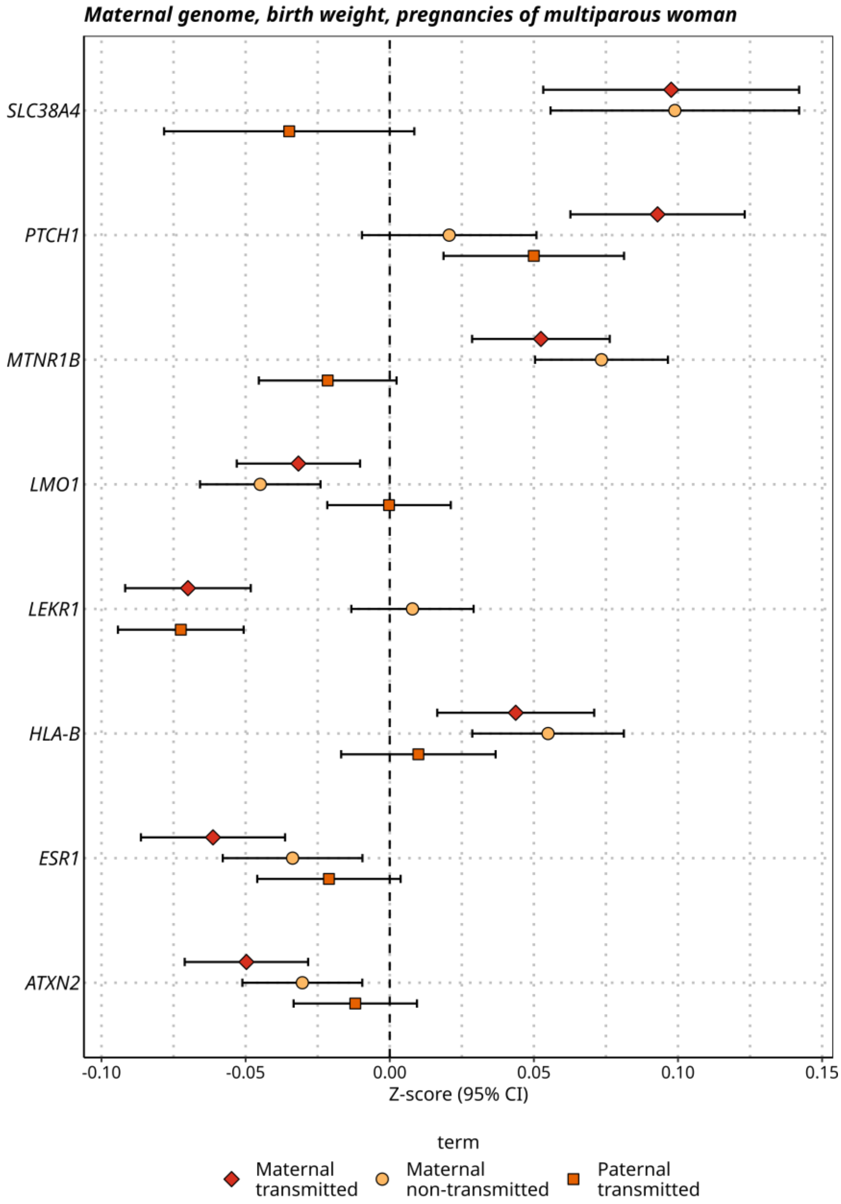
Maternal genome, birth weight, pregnancies of multiparous woman
SLC38A4
PTCH1
MTNR1B
LMO1
LEKR1
HLA-B
ESR1
ATXN2
-0.10
-0.05
0.00
0.05
0.10
0.15
Z-score (95% CI)
term
Maternal transmitted
Maternal non-transmitted
Paternal transmitted

**C**

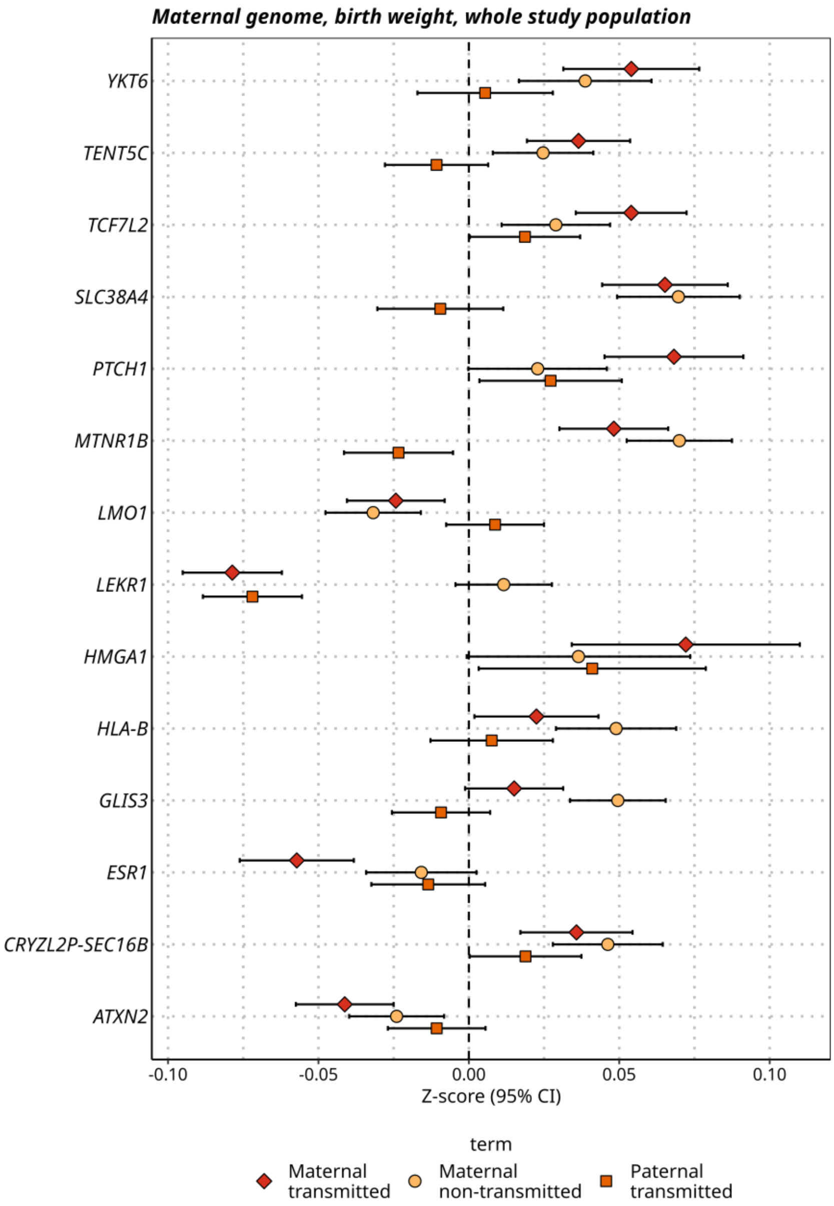


**Fig. S8.** The effect origin using mother-child duos for SNPs reaching the genome wide significance threshold in (A) birth weight maternal genome pregnancies of nulliparous woman ($n_{duos}$=21,789), (B) birth weight maternal genome pregnancies of multiparous woman ($n_{duos}$=28,690), (C) birth weight maternal genome whole study population ($n_{duos}$=50,479). Variants are displayed as the nearest protein-coding gene on the y-axis.

A

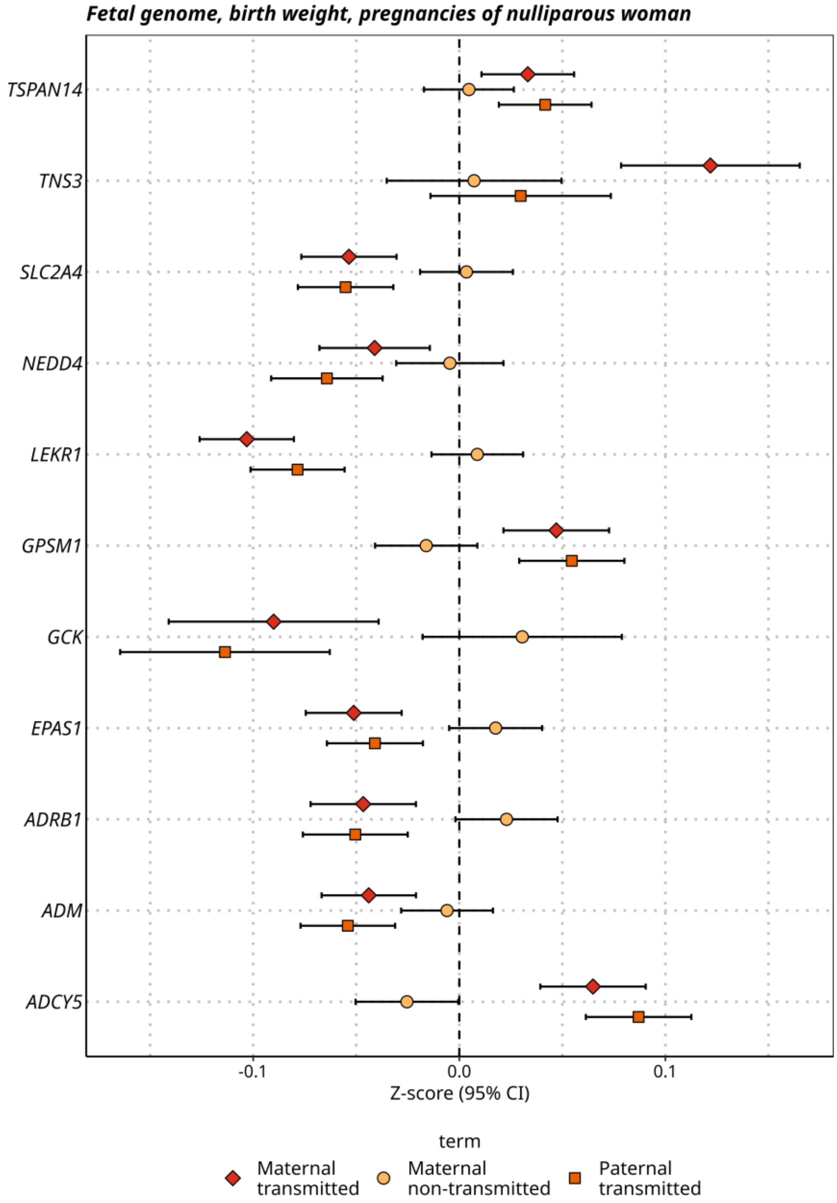

Fetal genome, birth weight, pregnancies of nulliparous woman
TSPAN14
TNS3
SLC2A4
NEDD4
LEKR1
GPSM1
GCK
EPAS1
ADRB1
ADM
ADCY5
-0.1
0.0
0.1
Z-score (95% CI)
term
Maternal transmitted
Maternal non-transmitted
Paternal transmitted

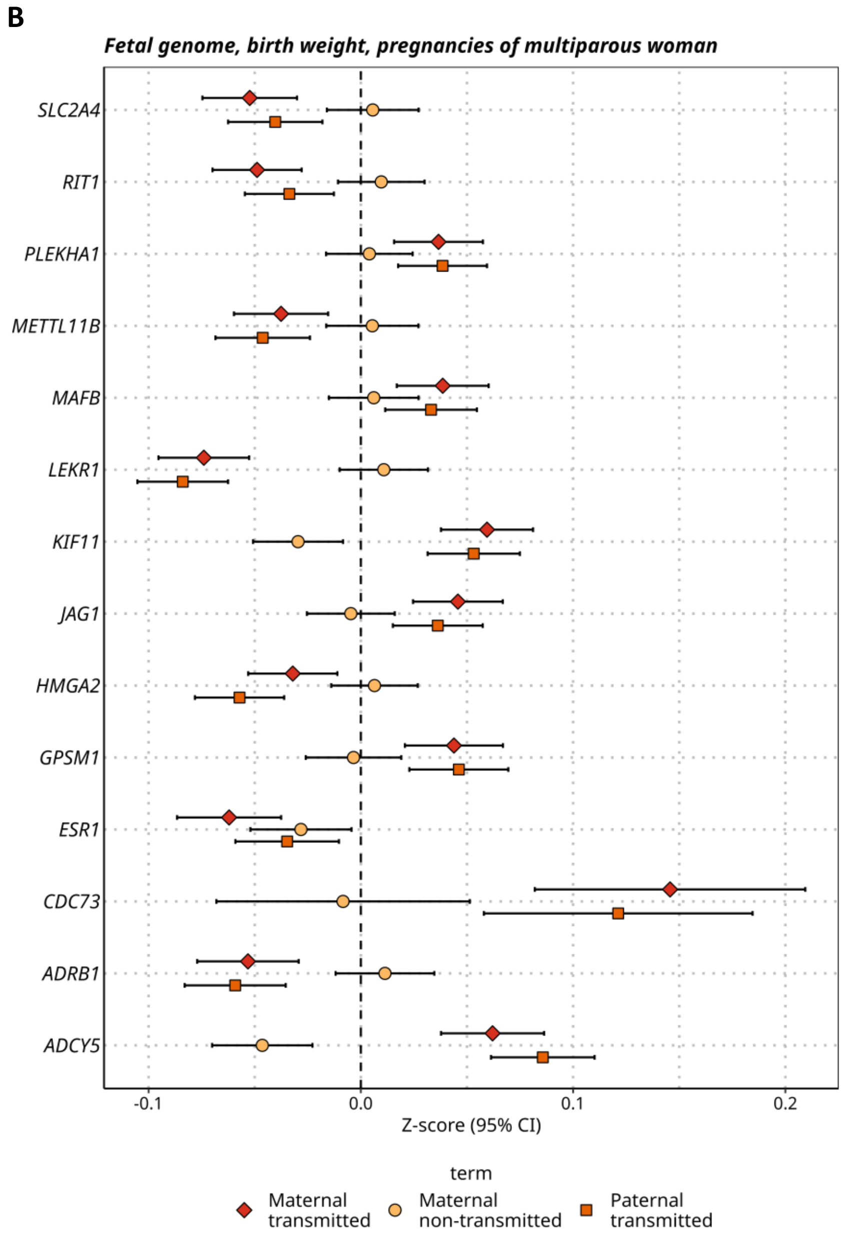
B
Fetal genome, birth weight, pregnancies of multiparous woman
SLC2A4
RIT1
PLEKHA1
METTL11B
MAFB
LEKR1
KIF11
JAG1
HMGA2
GPSM1
ESR1
CDC73
ADRB1
ADCY5
-0.1
0.0
0.1
0.2
Z-score (95% CI)
term
Maternal transmitted
Maternal non-transmitted
Paternal transmitted

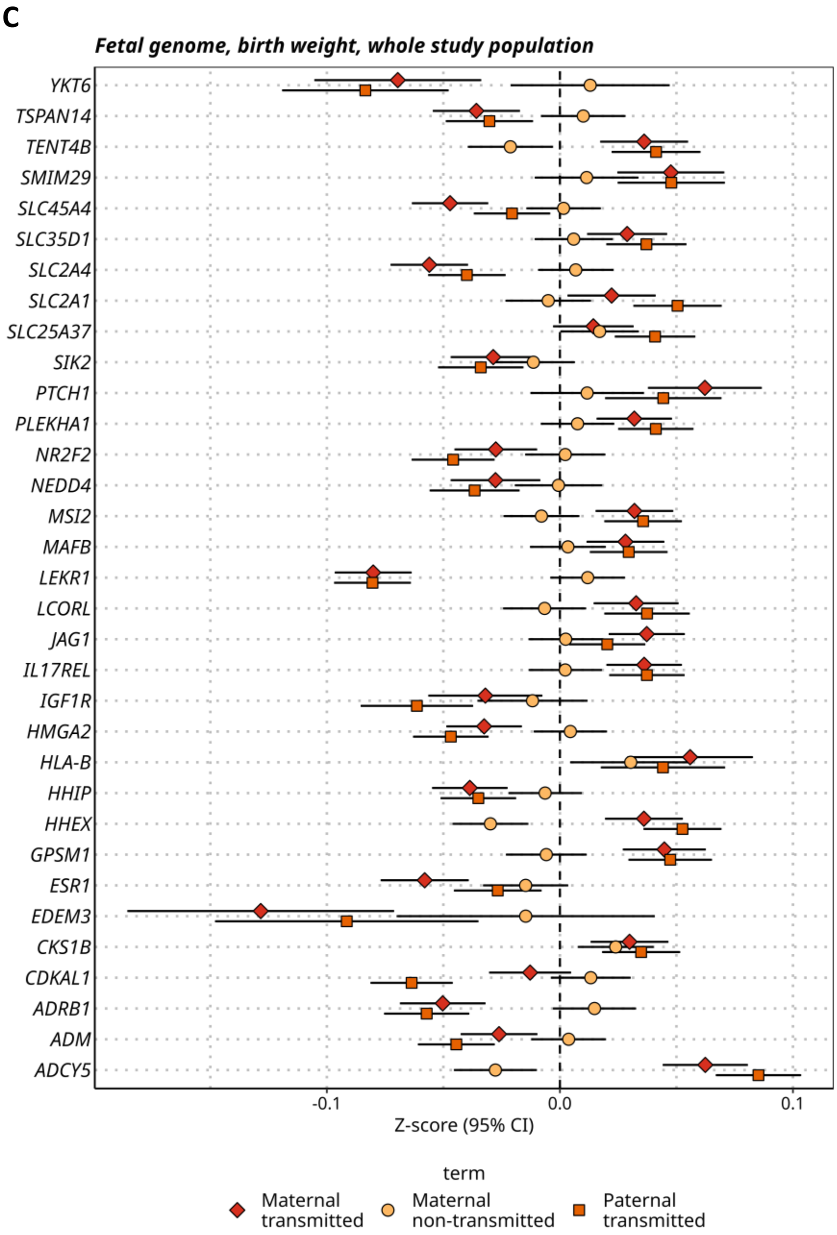


**Fig. S9.** The effect origin using mother-child duos for SNPs reaching the genome wide significance threshold in (A) birth weight fetal genome pregnancies of nulliparous woman ($n_{duos}$=21,779), (B) birth weight fetal genome pregnancies of multiparous woman ($n_{duos}$=28,016), (C) birth weight fetal genome whole study population ($n_{duos}$=49,795). Variants are displayed as the nearest protein-coding gene on the y-axis.

A

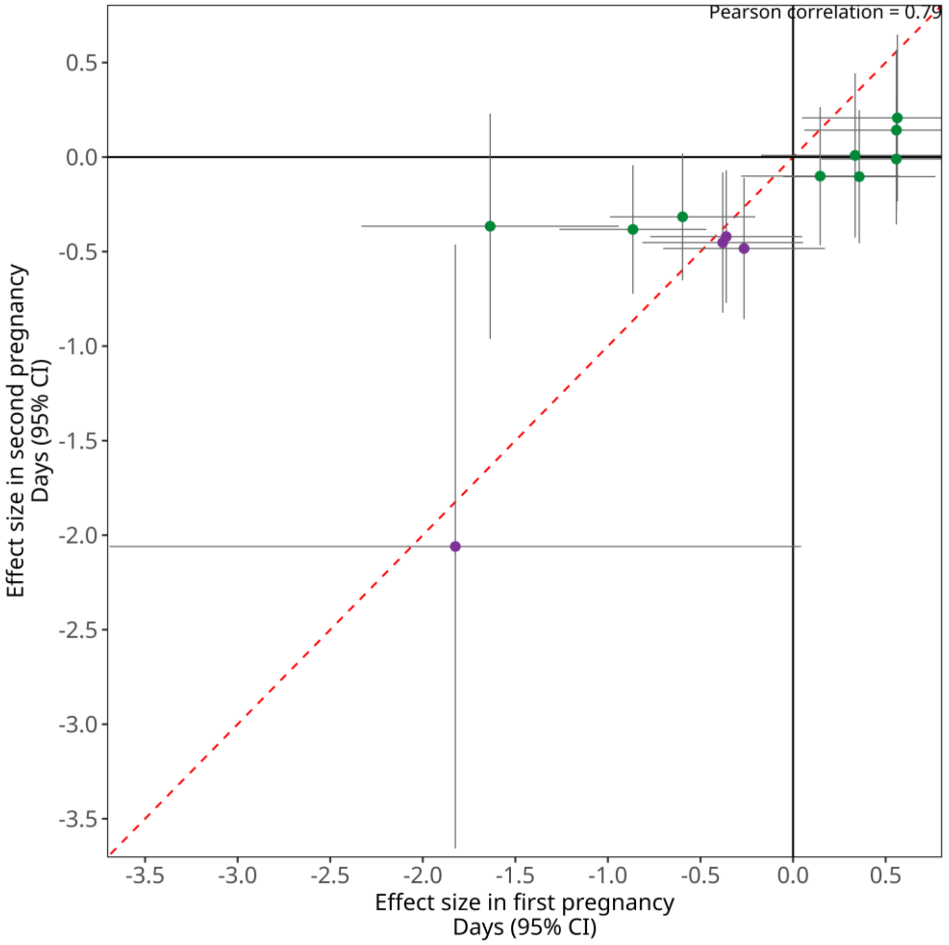

Pearson correlation = 0.79
0.5
0.0
-0.5
-1.0
-1.5
-2.0
-2.5
-3.0
-3.5
Effect size in second pregnancy
Days (95% CI)
-3.5
-3.0
-2.5
-2.0
-1.5
-1.0
-0.5
0.0
0.5
Effect size in first pregnancy
Days (95% CI)

**B**

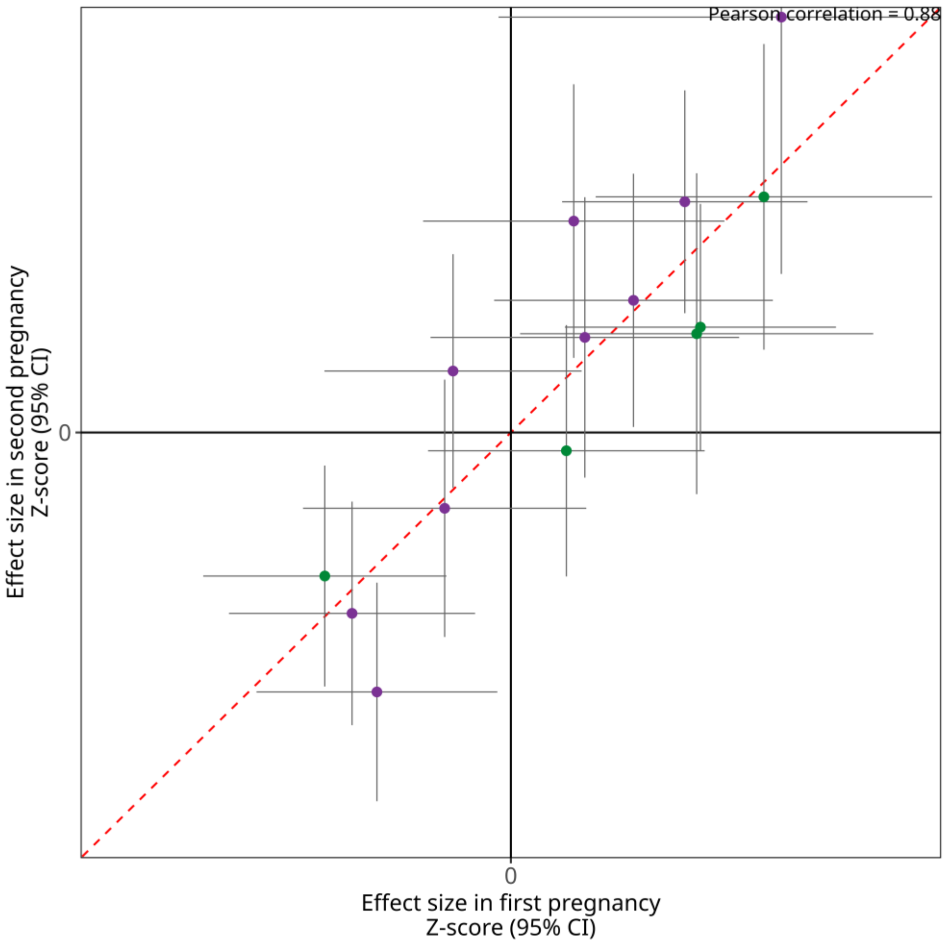


**Fig. S10.** Effect sizes of significant loci in (A) gestational duration (days) and (B) birth weight (z-score) in the first pregnancy and second pregnancy. The figure only includes independent variants that reached genome-wide significance for gestational duration on maternal genome or for birth weight on maternal genome across all parity groups. Estimates with a larger absolute effect in the first pregnancy are coloured in green, while estimates with a larger absolute effect in the second pregnancy are coloured in purple. The estimates are based on mothers with two pregnancies in MoBa (first and second pregnancy, n=5,895 for gestational duration and n=6,942 for birth weight). The red dashed line is the identity line. The error bars are the 95% confidence interval. The Pearson correlation between the effect sizes in first and second pregnancy is displayed at the top.

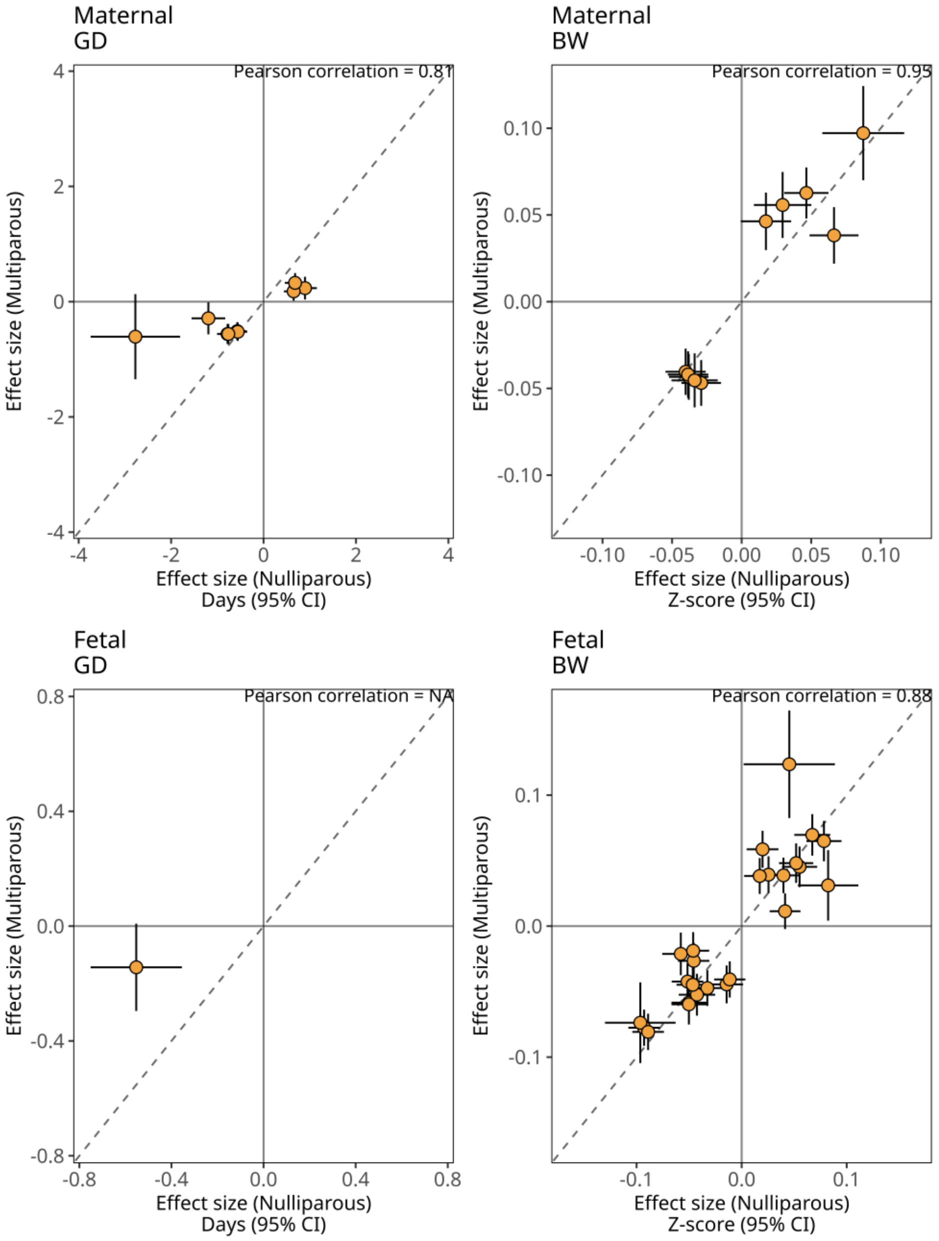


**Fig. S11.** Effect sizes (differences in days or z-score) for gestational duration (GD) and birth weight (BW) in the maternal (top) and fetal (bottom) parity stratified genome-wide association studies. The figure only includes variants that reached genome-wide significance in either the nulliparity GWAS or multiparity GWAS. The dashed line is the identity line. The error bars are the 95% confidence interval. The Pearson correlation between the parity-stratified genetic effects for each specific outcome is displayed at the top of each subfigure.

**A**

Maternal genome

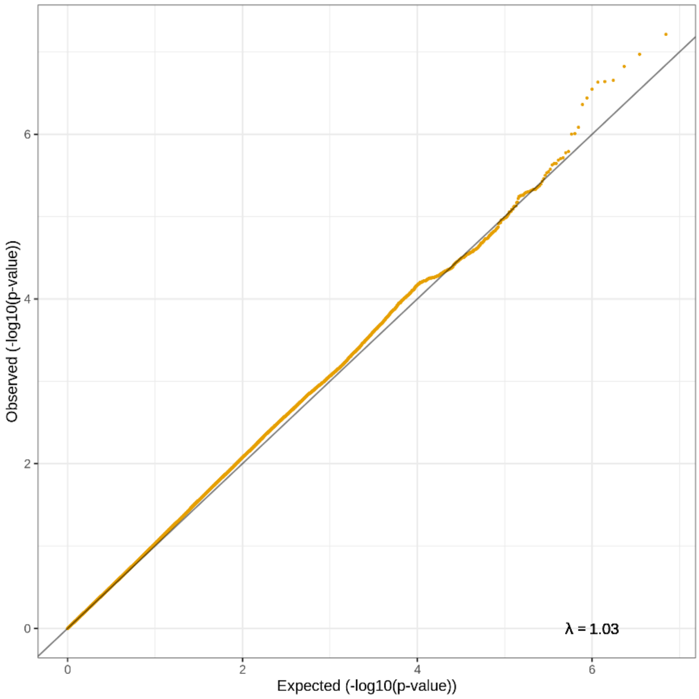


**B**

Fetal genome

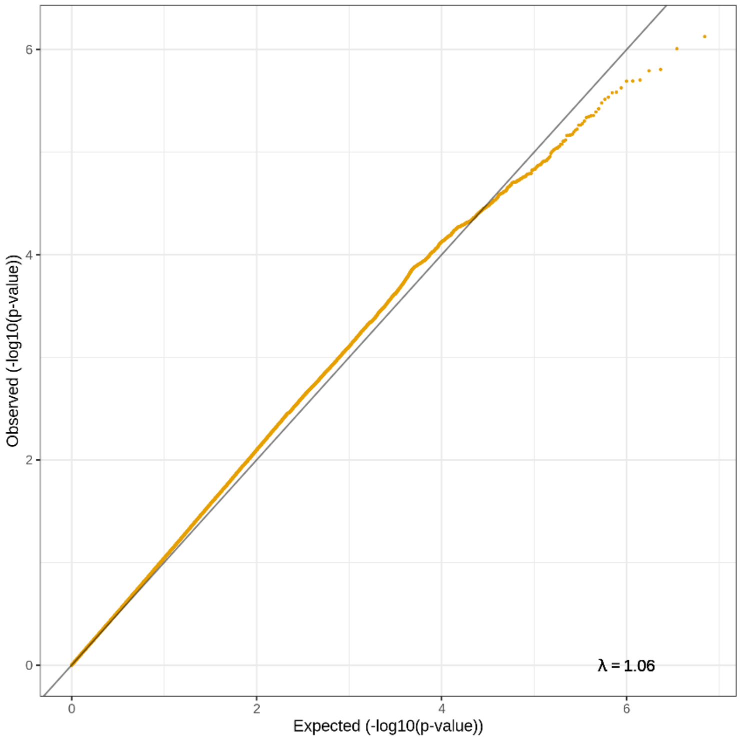

**C**

Maternal genome

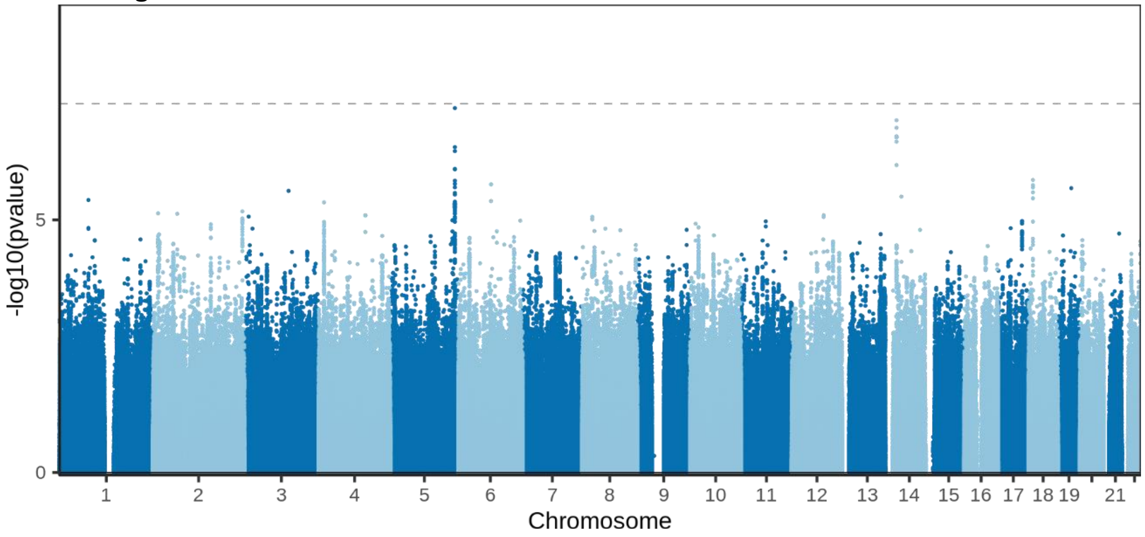


**D**

Fetal genome

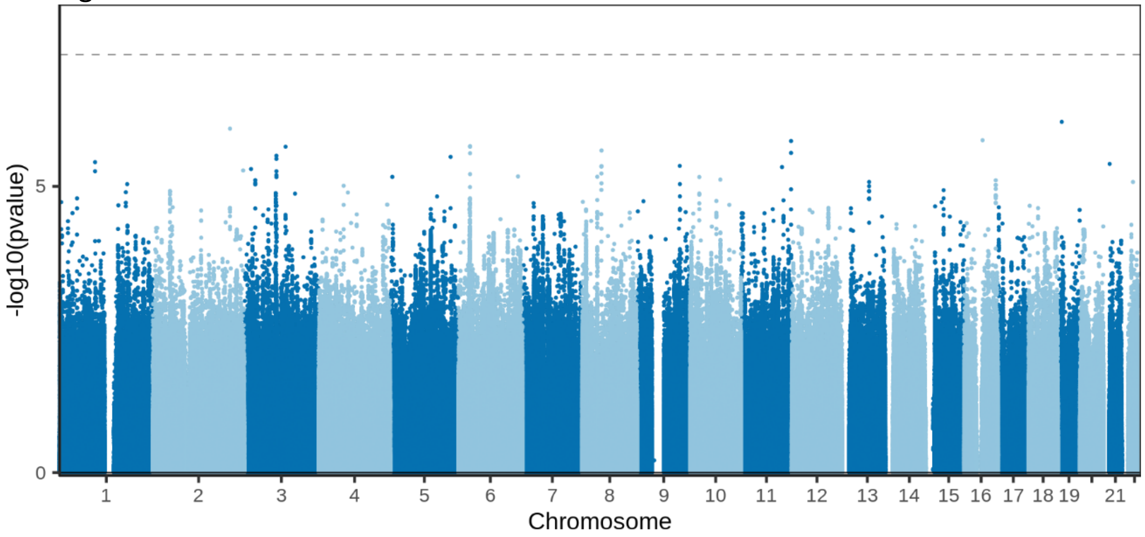


**Fig. S12.** QQ-plots and Manhattan plots for the genome-wide association study between first and later-born (as a case-control study). The QQ-plots shown in (A) for the maternal genome and (B) for the fetal genome include the lambda genomic control and demonstrate little to no test statistic inflation. In the Manhattan plots for (C) the maternal genome and (D) the fetal genome, the dashed line represents the genome-wide significant level (p-value = $5 \times 10^{-8}$). The x-axis shows the chromosome position and y-axis the two-sided p-value of the GWAS.

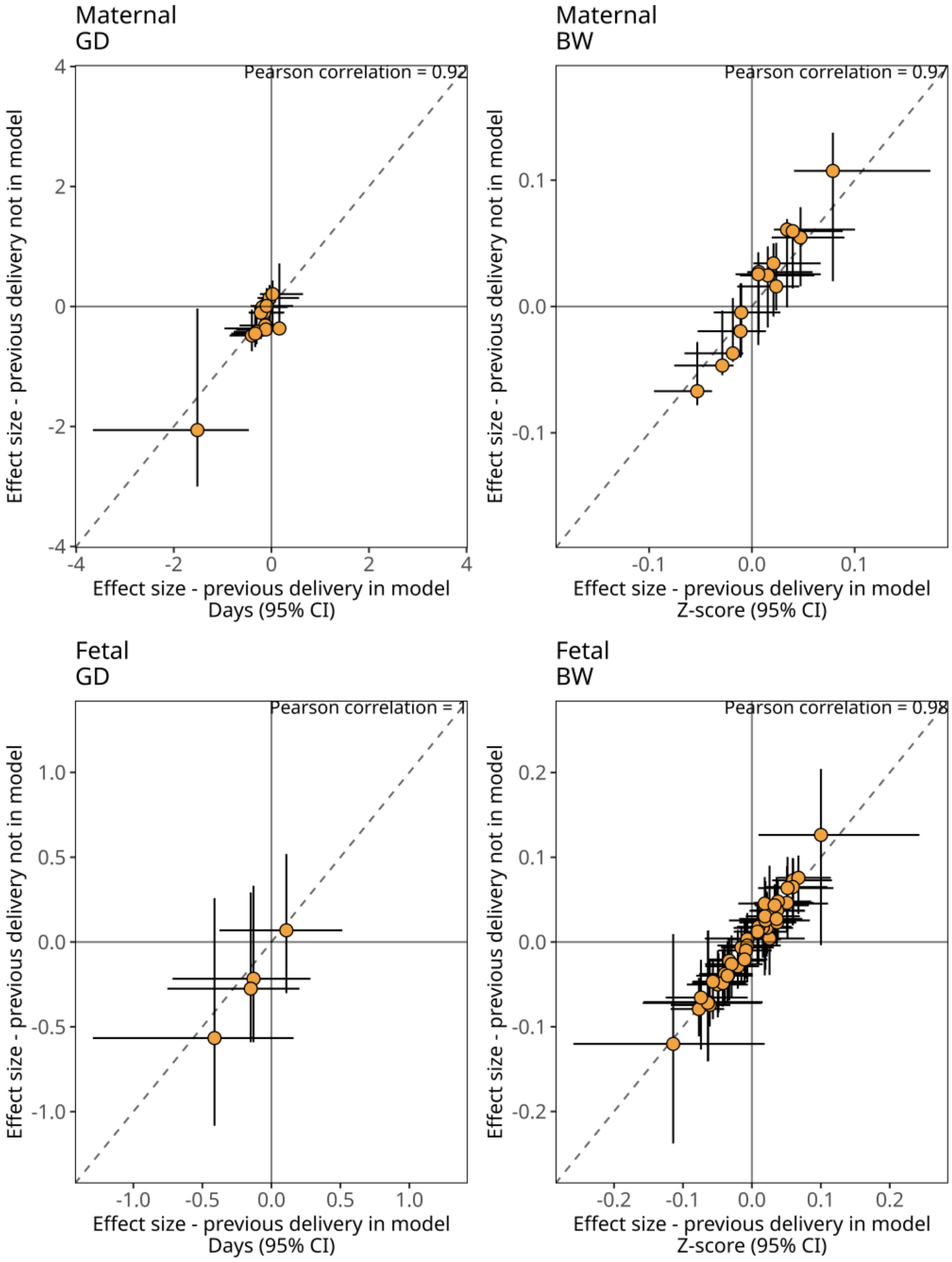


**Fig. S13.** Effect sizes (differences in days for gestational duration, GD, or z-score for birth weight, BW) for the second pregnancy in the maternal genome (top) and fetal genome (bottom). The figure only includes variants that reached genome-wide significance for each phenotype and genome in this study (across all parity groups). The estimates are based on mothers with two pregnancies in MoBa (first and second pregnancy; $n_{\text{gestational duration, maternal genome}}$=5,896, $n_{\text{birth weight, maternal genome}}$=6,942, $n_{\text{gestational duration, fetal genome}}$=3,021 and $n_{\text{birth weight, fetal genome}}$=3,872). The y-axis shows the effects of the main linear analysis. The x-axis shows estimates from corresponding linear models additionally adjusting for the outcome of the first pregnancy (GD in days or BW in z-scores). The dashed line is the identity line. The error bars are the 95% confidence interval. The Pearson correlation is displayed at the top of each subfigure.

**A**

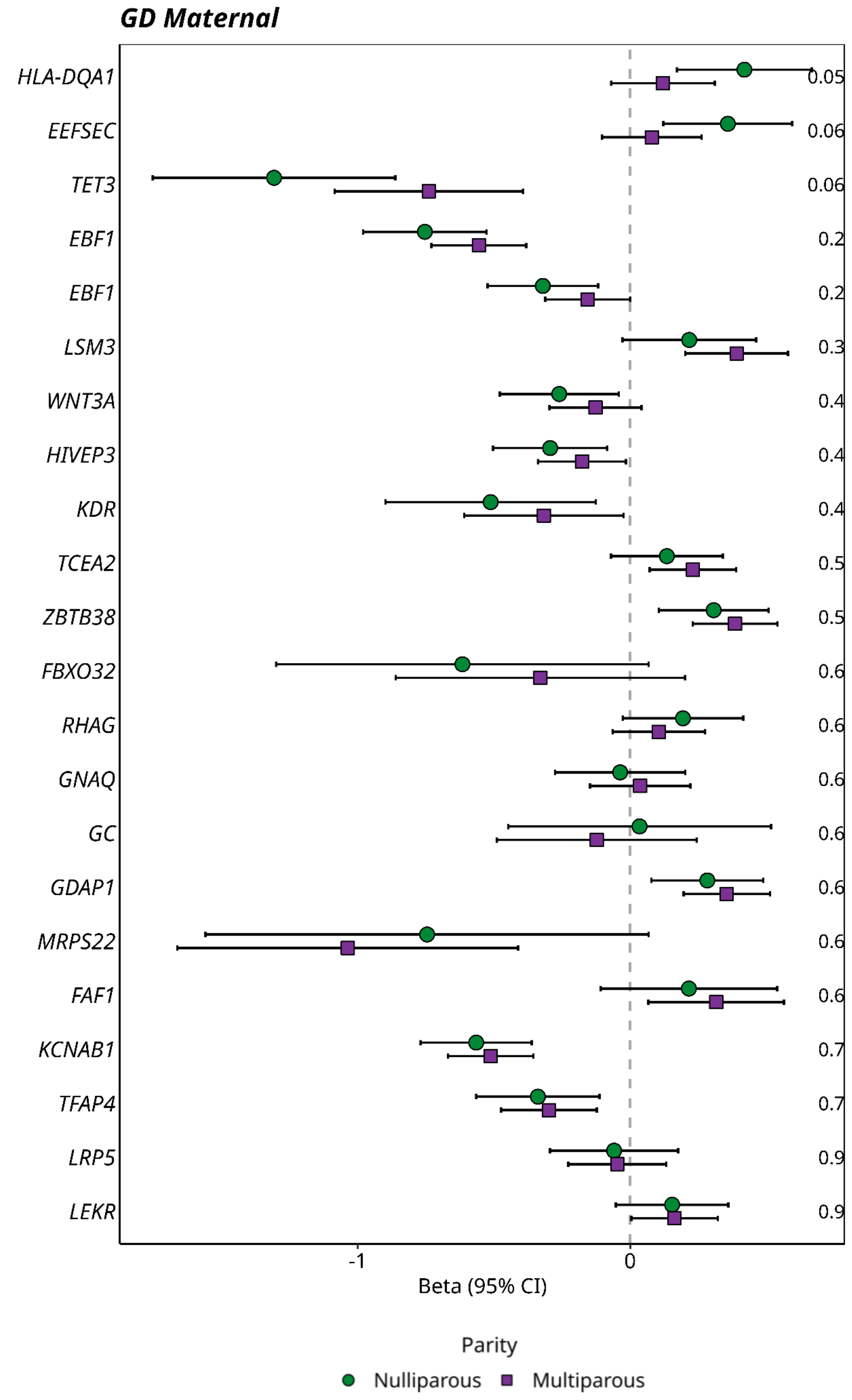
GD Maternal
HLA-DQA1
EEFSEC
TET3
EBF1
EBF1
LSM3
WNT3A
HIVEP3
KDR
TCEA2
ZBTB38
FBXO32
RHAG
GNAQ
GC
GDAP1
MRPS22
FAF1
KCNAB1
TFAP4
LRP5
LEKR
0.05
0.06
0.06
0.2
0.2
0.3
0.4
0.4
0.4
0.5
0.5
0.6
0.6
0.6
0.6
0.6
0.6
0.6
0.7
0.7
0.9
0.9
-1
0
Beta (95% CI)
Parity
Nulliparous
Multiparous

**B**

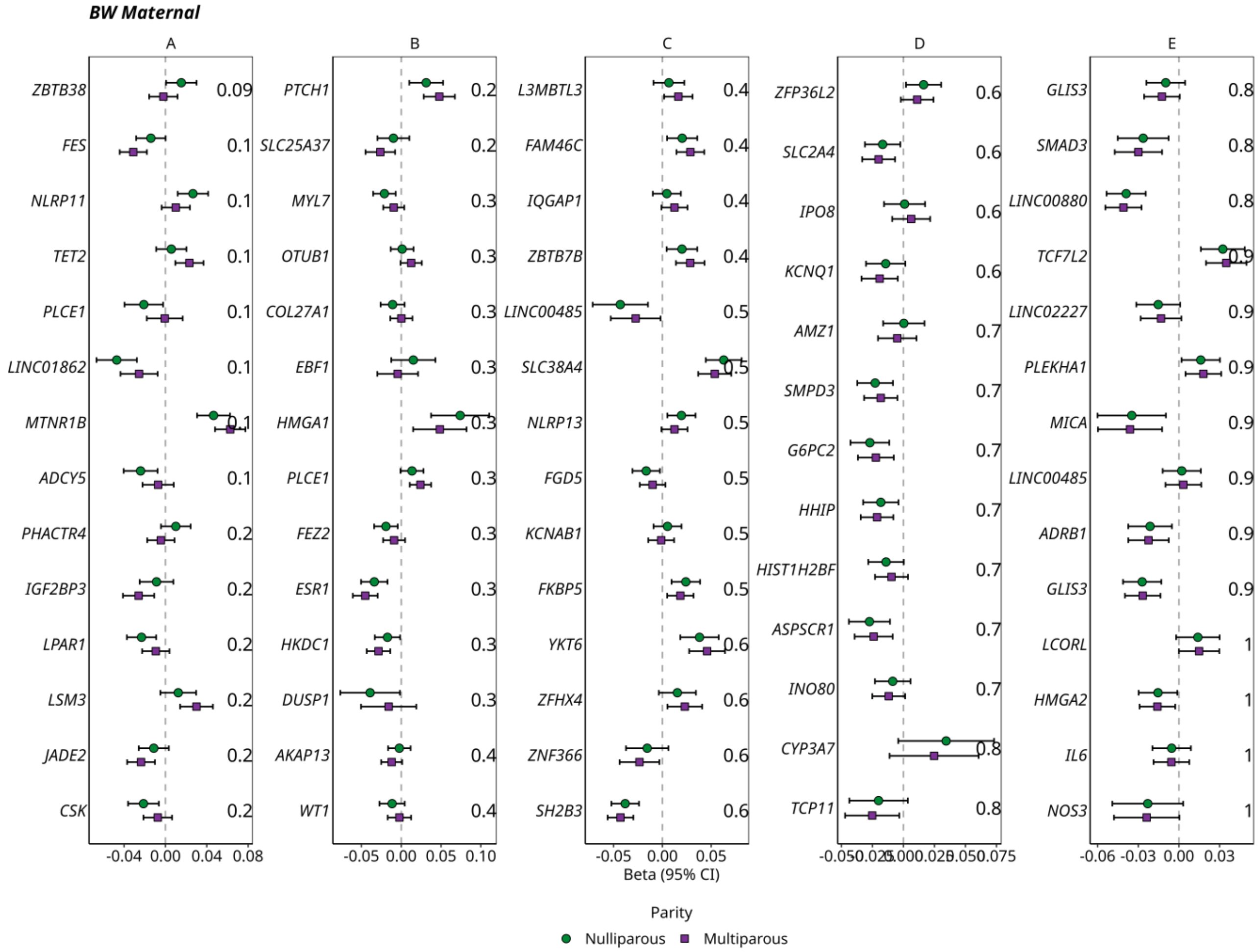

BW Maternal
A
B
C
D
E
ZBTB38
FES
NLRP11
TET2
PLCE1
LINC01862
MTNR1B
ADCY5
PHACTR4
IGF2BP3
LPAR1
LSM3
JADE2
CSK
0.09
0.1
0.2
PTCH1
SLC25A37
MYL7
OTUB1
COL27A1
EBF1
HMGA1
PLCE1
FEZ2
ESR1
HKDC1
DUSP1
AKAP13
WT1
0.3
0.4
L3MBTL3
FAM46C
IQGAP1
ZBTB7B
LINC00485
SLC38A4
NLRP13
FGD5
KCNAB1
FKBP5
YKT6
ZFHX4
ZNF366
SH2B3
0.5
0.6
ZFP36L2
SLC2A4
IPO8
KCNQ1
AMZ1
SMPD3
G6PC2
HHIP
HIST1H2BF
ASPSCR1
INO80
CYP3A7
TCP11
0.7
0.8
GLIS3
SMAD3
LINC00880
TCF7L2
LINC02227
PLEKHA1
MICA
LINC00485
ADRB1
GLIS3
LCORL
HMGA2
IL6
NOS3
0.9
1
-0.04 0.00 0.04 0.08
-0.05 0.00 0.05 0.10
-0.05 0.00 0.05
-0.050 -0.025 0.000 0.025 0.050 0.075
-0.06 -0.03 0.00 0.03
Beta (95% CI)
Parity
Nulliparous
Multiparous

**C**

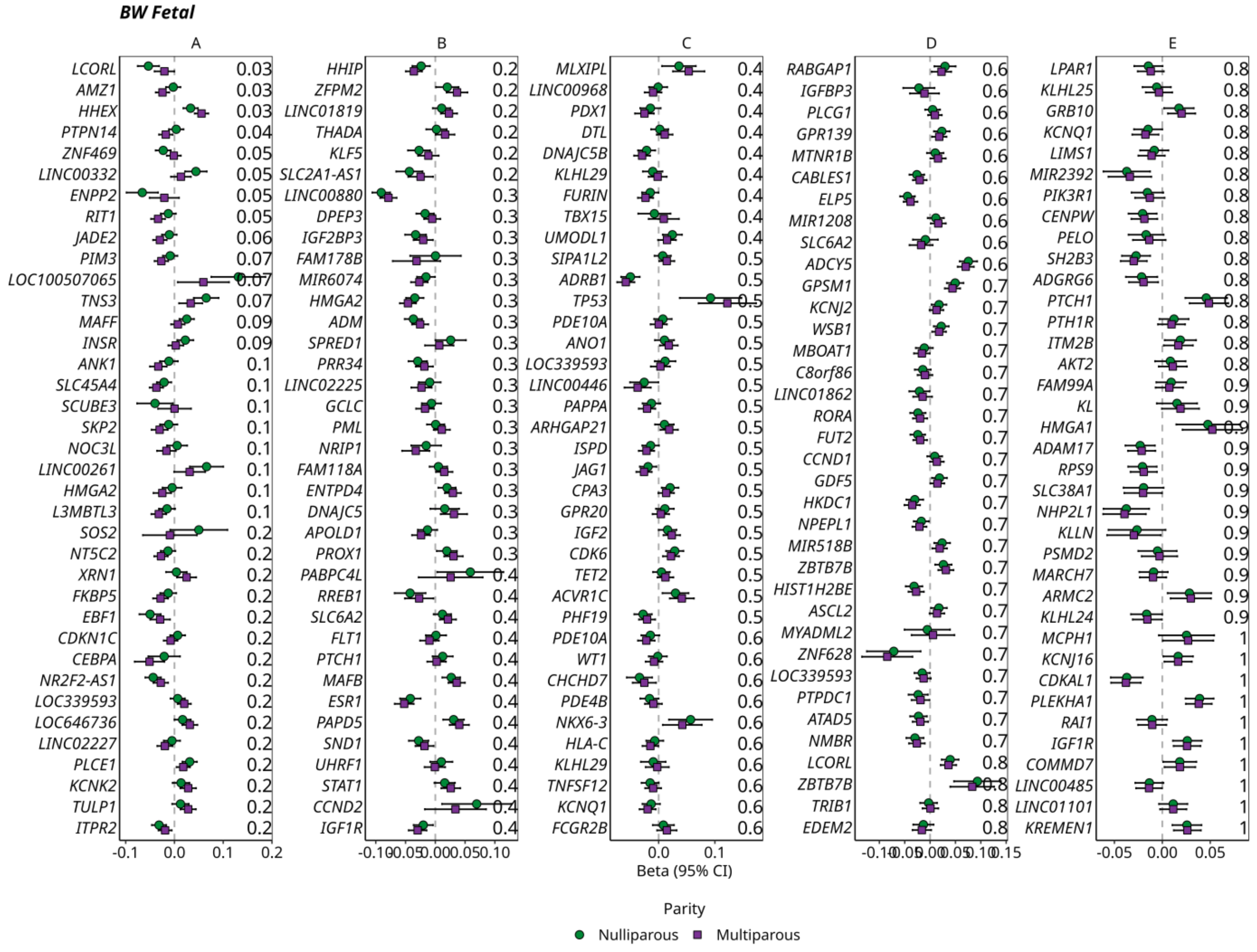


**Fig. S14.** Parity stratified effect sizes (n ~ 28,000 mothers/ children) for (A) gestational duration (GD) maternal genome, (B) birth weight (BW) maternal genome and (C) BW fetal genome. The green circle are the effect sizes in pregnancies of nulliparous women and the purple square are the effect sizes in pregnancies multiparous women. The error bars are the 95% confidence interval. Variants are displayed as the nearest protein-coding gene on the y-axis. The interaction p-value for each variant is displayed to the right. Only variants previously associated with the specific outcome in were included. The five variants with the lowest interaction p-value are illustrated in Fig. 2.

**A**

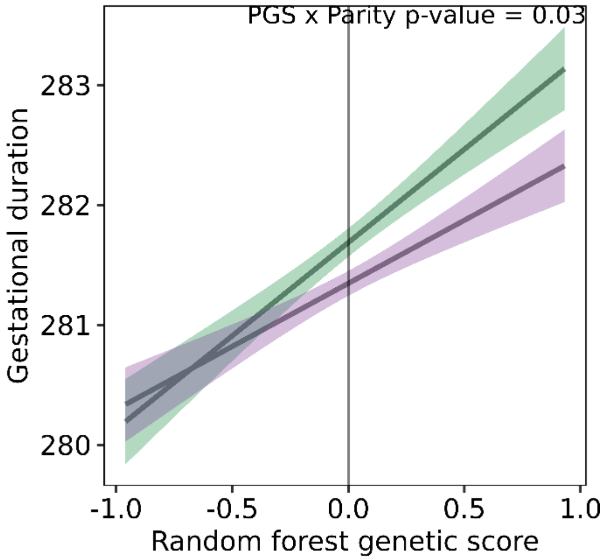


**B**

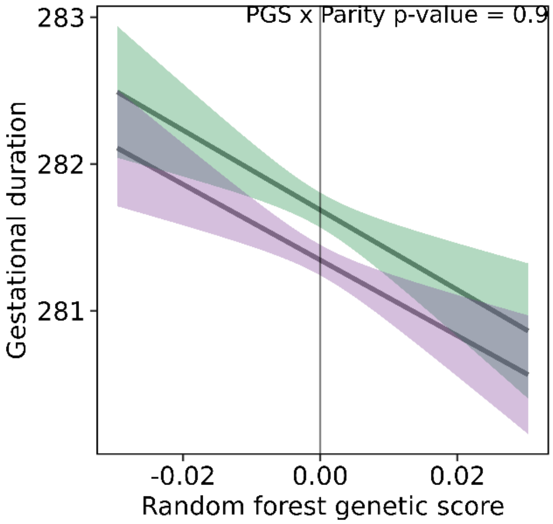


**Fig. S15.** The interaction between the maternal random forest genetic score and nulliparity on gestational duration using (A) the MnT allele and (B) the PT allele. The slopes highlight the effect of the random forest genetic score on gestational duration by parity and are based on the estimates from the linear model used when testing the interaction ($n_{nulliparity}$=17,739 and $n_{multiparity}$=23,097, Table S13). The x-axis shows the maternal random forest genetic score for Mnt and PT allele, respectively, and the y-axis the gestational duration. The interaction p-value is displayed in the top right of the figure.

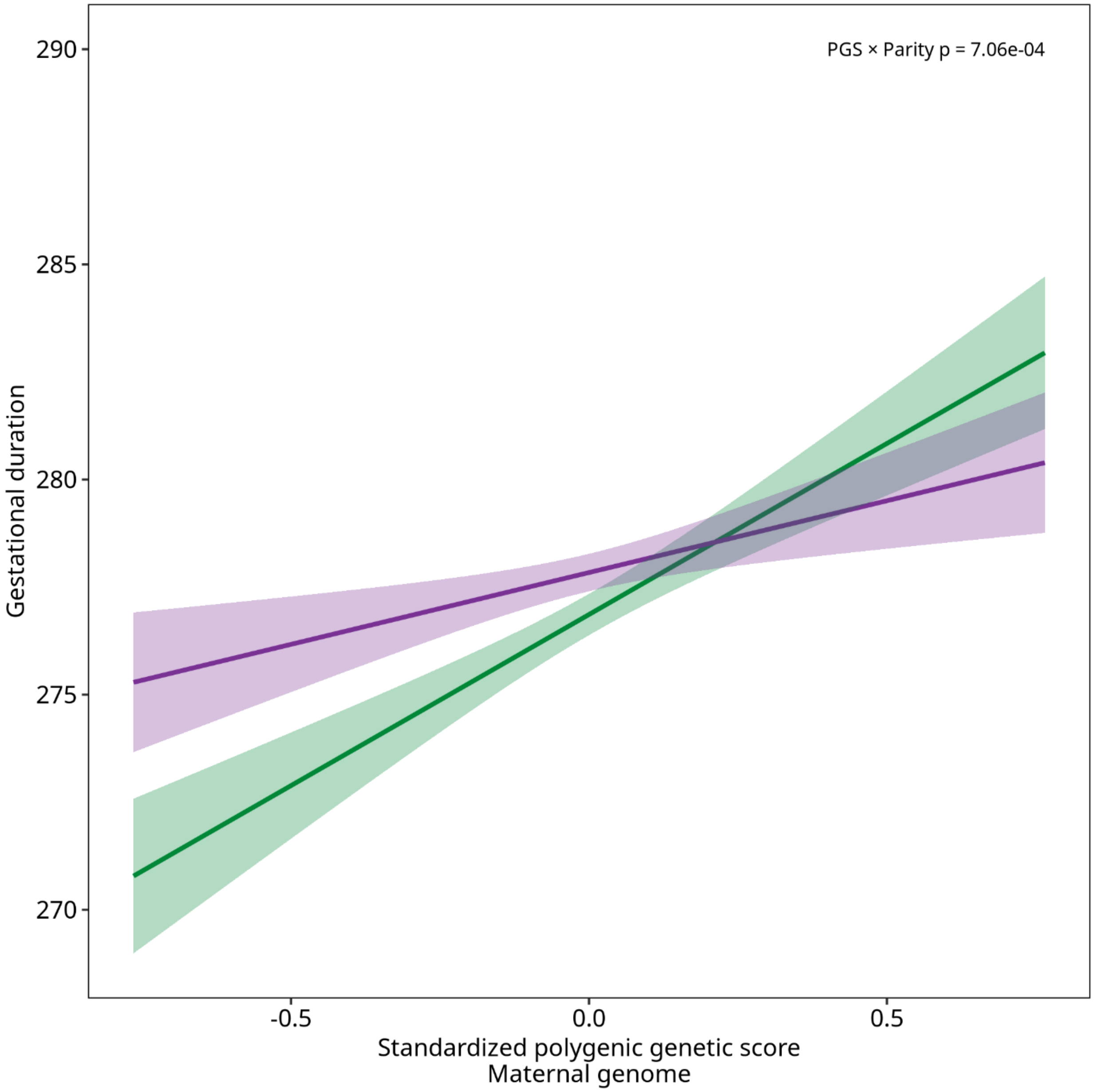


**Fig. S16.** The interaction between the genome-wide polygenic score (PGS) of gestational duration on the maternal genome and nulliparity. The PGS was constructed with weights from the current MoBa GWAS (whole study population) and tested with the ALSPAC cohort. The slopes highlight the effect of the PGS on gestational duration by parity (green line: nulliparity (n= 2,855), purple line: multiparity (n= 3,468)) and are based on the estimates from the linear model used when testing the interaction (Table S13). The x-axis shows the PGS and the y-axis shows the gestational duration. The interaction p-value is displayed in the top right of the figure.

**Supplementary table captions**

**Table S1.** Sample size and characteristics for each phenotype and genome. The sample size shown is the maximum sample size for each phenotype and genome.

**Table S2.** Summary statistics from the genome-wide significant SNPs (p-value < 5e-8) for genome-wide association studies (maternal genome) on gestational duration (additive model).

**Table S3.** Summary statistics for variants that associate with either gestational duration or offspring birth weight in maternal or fetal genome in following previous studies (2-6). Some of the previously identified variants associated with gestational duration or birth weight did not pass quality control or were not part of the imputation panel. If a variant within the genomic region had an r2 > 0.3 to the missing top SNP, this SNP was used as a proxy. However, some proxies are missing for some variants and the X chromosome is not included in this study; these entries are marked with "-".

**Table S4**. Haplotype analysis to resolve effect origin using the model: outcome = β0 + β1 MT + β2 MnT + β3 PT + β4 covariate4 + ... + ε. The terms h1, h2 and h3 stand for maternal transmitted (MT), maternal non-transmitted (MnT) and paternal transmitted (PT), respectively. A variant was classified as having a maternal-only effect when PT 95% CI overlapped zero and the MnT 95% CI did not overlap zero. A fetal-only effect was defined when MnT 95% CI overlapped zero and the PT 95% CI did not overlap zero, or when only MT 95% CI did not overlap zero. Variants were classified as having both maternal and fetal effects when the 95% CI for both PT and MnT did not overlap zero. These models were run using data from 17,869 up to 50,47 mother-child duos across outcomes, genotypes, and parity groups.

**Table S5.** Summary statistics from the genome-wide significant SNPs (p-value < 5e-8) for genome-wide association studies (fetal genome) on gestational duration (additive model).

**Table S6.** Summary statistics from the genome-wide significant SNPs (p-value < 5e-8) for genome-wide association studies (maternal genome) on birth weight (z-score) (additive model).

**Table S7.** Summary statistics from the genome-wide significant SNPs (p-value < 5e-8) for genome-wide association studies (fetal genome) on birth weight (z-score) (additive model).

**Table S8.** Sample size and characteristics for each of the phenotype and genome in ALSPAC. The sample size shown is the maximum sample size for each phenotype and genome.

**Table S9.** Replication analyses of the loci reaching the genome-wide significant threshold in study using the Avon Longitudinal Study of Parents and Children (ALSPAC) cohort.

**Table S10.** Interactions between previously associated SNPs (2-6) and its respective outcome (gestational duration or birth weight) for each genome (maternal and fetal) in first pregnancies and subsequent ones. P-value refers to one for the interaction term SNP x parity (using whole population). Effect size, standard error, and confidence interval were obtained after fitting a linear regression split by parity.

**Table S11.** The Bonferroni correction, adjusting for the total number of variants analysed for each outcome and genome.

**Table S12.** Disentangle the direct effects of parity from the joint distribution with maternal age in previous associated SNP.

**Table S13.** The estimated betas (β) and p-value of the linear model used to test for random forest genetic scores x nulliparity for all phenotypes and genomes (only using the dosage of genetic variants to create the random forest genetic scores) in MoBa; Phenotype = $\beta 0 + \beta 1$ random forest genetic score + $\beta 2$ nulliparity + $\beta 3$ random forest genetic score*nulliparity, the random forest genetic score was scaled so its mean was zero. For the ALSPAC replication, the following model was used: gestational duration = $\beta 0 + \beta 1$ PRS-CS genetic score maternal + $\beta 2$ PRS-CS genetic score fetal + $\beta 3$ nulliparity + $\beta 4$ PRS-CS genetic score maternal*nulliparity + $\beta 5$ PRS-CS genetic score fetal*nulliparity (here using a genome-wide polygenic score (PGS) of gestational duration).

**Table S14.** The estimated betas (β) and p-value of the linear model used to test for random forest genetic scores x nulliparity for all phenotypes and genomes (using both the dosage of genetic variants and environmental factors to create the random forest genetic scores). (Phenotype = $\beta 0$ + $\beta 1$ random forest genetic score + $\beta 2$ nulliparity + $\beta 3$ random forest genetic score*nulliparity, the random forest genetic score was scaled so its mean was zero).

**Table S15.** Prediction accuracy ($r^2$) of each phenotype, genome and population (parity stratified or not) in random forest models.

**Table S16.** Associations between the PGS of gestational duration derived from nulliparous and multiparous women (from MoBa) on gestational duration in nulliparous and multiparous women from ALSPAC.

**Table S17.** SNP heritability for gestational duration and birth weight in the maternal and foetal genomes by parity (n ~ 28,000 mothers/ children).

**Table S18.** Number of significant variants based on sample size.
